# Monitoring the propagation of COVID-19-pandemic first waves

**DOI:** 10.1101/2020.05.09.20096768

**Authors:** William Knafo

**Affiliations:** Laboratoire National des Champs Magnétiques Intenses, CNRS-UPS-INSA-UGA, 143 Avenue de Rangueil, 31400 TOULOUSE, FRANCE

## Abstract

A phenomenological approach is proposed to monitor the propagation of the COVID-19-pandemic first waves. A large set of data collected at a worldwide scale during the first months of 2020 is compiled into series of semi-logarithmic plots, for a selection of thirty-two countries from the five continents. Three regimes are identified in the propagation of an epidemic wave: a pre-epidemic regime 1, an exponential-growth regime 2, and a resorption regime 3. A two-parameters scaling of the first-wave death variation reported in China is used to fit those reported in other countries. Comparison is made between the propagation of the pandemic in different countries, which are classified in four groups, from group A where the pandemic first waves were contained efficiently, to group D where the pandemic first waves widely spread. Group A is mainly composed of Asian countries, where fast and efficient measures have been applied. Group D is composed of Western-Europe countries and the United States of America, where late decisions and confused political communication (pandemic seriousness, protection masks, herd immunity etc.) led to significant death tolls. The threat of large resurging epidemic waves after a hasty lockdown lift is discussed, in particular for the countries from group D, where the number of contagious people remained high in the beginning of May 2020. The situation is opposite in Asian countries from group A, where the number of contagious people was successfully maintained to a low level. In particular, the results obtained by Hong Kong and South Korea are highlighted, and the measures taken there are presented as virtuous examples that other countries may follow.

## 1- Introduction

In the first months of 2020, the pandemic first-waves spread of the coronavirus disease 2019 (COVID-19) has affected most of the countries worldwide [1]. This disease, caused by severe acute respiratory syndrome coronavirus 2 (SARS-CoV-2), was first reported in end 2019 in Hubei province, China [2],[3]. At the time of writing this paper (9 May 2020), almost three hundred thousand deaths have been reported and multiple challenges have emerged: slowing down the spread of the virus, offering adapted medical cares, saving lives, developing a vaccine to immunize the population, and anticipating a forthcoming economic crisis. As first step, slowing down the pandemic propagation is essential to limit the number of deaths occurring in a few-week timescale and avoid a cascade of related issues. Without this, a wild and uncontrolled exponential propagation could lead to cumulative death tolls of up to one or a few percent of a population. This corresponds to a situation where herd immunity would be achieved in a ‘natural’ manner [4],[5],[6]. The target to avoid such situation offers a rare case where scientists can directly guide politicians and where their recommendations on short- and mid-term decisions can have enormous impacts for the community. They can monitor the pandemic statistics, they can model it, they can propose solutions to slow down the pandemic propagation, they can follow or anticipate the impacts of given series of political decisions. These last weeks, several epidemiological models and reports emerged [7],[8],[9],[10],[11],[12],[13],[14],[15] some of them having impact in national press and immediate consequences on political decisions. In addition to the work from epidemiologists, modelling and graphical tools have been proposed by physicists (see for instance [16],[17],[18],[19]). In particular, phenomenological approaches, as that proposed in this work, are suited to monitor the propagation of a pandemic.

Here, a battery of semi-logarithmic plots on the propagation of the COVID-19 pandemic is given, for a selection of thirty-two countries from the different continents. A two-parameter scaling of the death data reported in China is used to fit the first epidemic waves in a selection of countries, where the spread was well-advanced in beginning of May 2020 (USA, Spain, Italy, United Kingdom, France and Germany). The presented graphs constitute simple tools to identify the trends and key moments in the propagation of the pandemic in a country. They offer an easy way to judge and compare the efficiency of social-distancing, containment and lock-down measures. The success of the measures taken in Asian countries is emphasized. The situation in several Western Europe countries and in the United States of America is opposite. Confused political communication about i) the appreciation of the pandemic’ seriousness [20],[21], ii) recommendations to wear protection masks [22], and iii) the consequences of a herd immunity scenario [23], has been observed. In these countries, the delay in the application of strong measures led to tens of thousands of deaths after the first epidemic waves. An early lift of the lockdown may also lead to the resurgence of epidemic waves. The monitoring tools compiled here, once updated, will help in forecasting resurging waves of the pandemic. The fatality rate and the question of achieving herd immunity, as well as the exponential consequences of a delay or inefficiency in the application of measures are discussed.

## 2- Materials and methods

Data presented here were extracted from the John Hopkins University [24] and Santé Publique France databases [25]. They were accessed on 9 May 2020 and correspond to confirmed cases and deaths tolls reported in thirty-two countries worldwide up to 8 May 2020.

## 3- Results

### 3.1- Spread of the pandemic in a selection of Asian and Western countries

Figure 1 presents the time variation of confirmed case and death tolls from a selection of six countries early hit by the COVID-19 pandemic, in a time scale covering fully or partly the first epidemic waves. The panels (a), (c), and (f) show the evolution from the 1 January to the beginning of May 2020 of the cumulative confirmed cases, cumulative deaths, and daily deaths in China, South Korea, Italy, Spain, France (mainland) and the United States of

**Figure 1:**
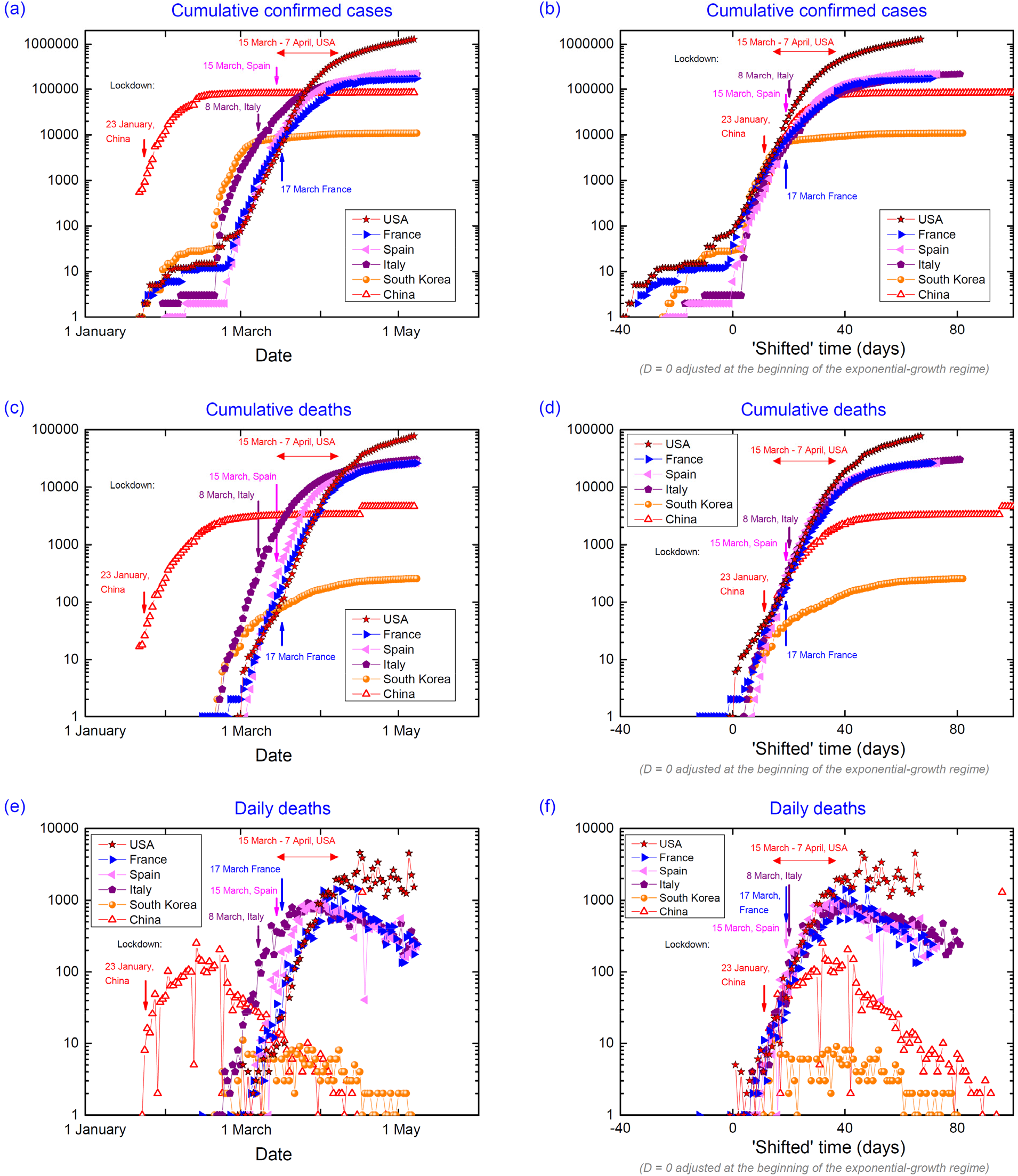
*Focus on six countries: China, South Korea, Italy, Spain, France (mainland) and the United States of America [26],[27],[28],[29],[30]*.

America. Time offsets between the variations from the different countries result from the delayed arrivals of the virus on their territory. China was the first country hit by the pandemic, whose first wave ended in late April 2020, with cumulative confirmed cases saturating at ≈ 80 000 and cumulated deaths saturating < 5 000 (initial saturation to ≈ 3 300 deaths corrected to ≈ 4600 on 17 April 2020). South Korea was hit a few weeks after China and was less affected than the other countries considered here, with < 11 000 cumulative confirmed cases and < 300 cumulative deaths in the beginning of May 2020. The United States of America were the last country of this selection to be hit, but they were the most affected with > 1 000000 cumulative confirmed cases and > 70 000 cumulative deaths in the beginning of May 2020. The three Western Europe countries were hit a few days after South Korea and were strongly affected, with ≈ 200 000 cumulative confirmed cases and ≈ 30 000 cumulated deaths in each country in the beginning of May 2020. These graphs show that, when the epidemic is active in a country, the number of cumulative deaths increases in an exponential manner, which leads to a linear increase in the semi-logarithmic scale of the graphs.

Complementarily graphs are shown in the panels (b,d,f) of Figure 1 where the cumulative confirmed cases, cumulative deaths and daily deaths are plotted as a function of a ‘shifted’ time. For each country, the ‘shifted’ time is adjusted so that the day *D* = 0 corresponds to the extrapolation to a number *N* = 1 of the exponential increase of cumulative deaths. A deviation from the exponential regime is observed in all countries a few days or weeks after the day *D* = 0. An epidemic peak corresponding to a several-week plateau in the time variation of the daily death number is visible for the six countries considered in Figure 1. This plateau was always maintained to < 10 daily deaths in South Korea. It reached 100 – 200 daily deaths in China, 500 – 1 000 daily deaths in the four considered Western Europe countries, and a maximum of 2 000 – 4 000 daily deaths in the United States of America.

Figure S1 in the Supplementary Materials generalizes the graphs from Figure 1 to a total of thirty-two countries worldwide. It confirms that all countries follow similar trajectories. A large scattering of the data is visible in the plots of the cumulative confirmed cases as a function of the ‘shifted’ time [Figure S1(c)]. Oppositely, the cumulative deaths plotted as a function of the ‘shifted’ time almost converge on a unique line in the exponential regime of the pandemic propagation [Figure S1(f)]. This difference can be explained as the number of cumulative confirmed cases is a less reliable quantity than the number of cumulated deaths, for the following reasons:

- The tests on the population are done with different financial resources, with different efforts, and different constancies, depending on the considered country. A large proportion of cases are not detected.
- When the number of cases increases, it becomes more difficult to detect all of them. Even in the countries equipped with the best detecting system, detection is less efficient when it approaches its maximal capacity (saturation of a detector).

In the next Sections, the number of deaths, thought to be more reliable, will be considered preferentially. We note that voluntary or involuntary failures in death counting can also occur. This was for instance the case in France, where uncomplete numbers of deaths (only deaths in hospitals) were communicated before 1 April 2020. In the United Kingdom, the deaths outside hospitals were also not counted before 29 April 2020. In many countries, only the deaths in hospitals have been counted so far.

Complementarily to Figure 1, Figure 2 presents a comparison, for a selection of nine countries in Asia and Western North hemisphere, of the variation with time of the daily confirmed cases and death tolls. For each country, the variation of the daily death number follows that of the daily confirmed cases, with a delay of 5-10 days. In China, a significant decrease of daily confirmed cases was observed 10 days after the setup of lockdown, and the epidemic peak in the number of daily deaths was observed 10 days later. After this peak, the number of daily deaths has decreased within an exponential decay, as indicated by the negative-slope linear variation in the semi-logarithmic scale of the graphs. Two months later, in the beginning of April, lockdown lift was decided. At this date, there were a few daily deaths and ≈ 50 – 100 daily confirmed cases in China. The case of South Korea is unique: after an early increase of daily confirmed cases, this number reached a maximum of ≈ 1 000 before strongly decreasing. In the end of April, less than 2 daily deaths and ≈ 10 daily confirmed cases were reported. Before May, the number of daily deaths has always been contained to less than 10 in South Korea.

**Figure 2:**
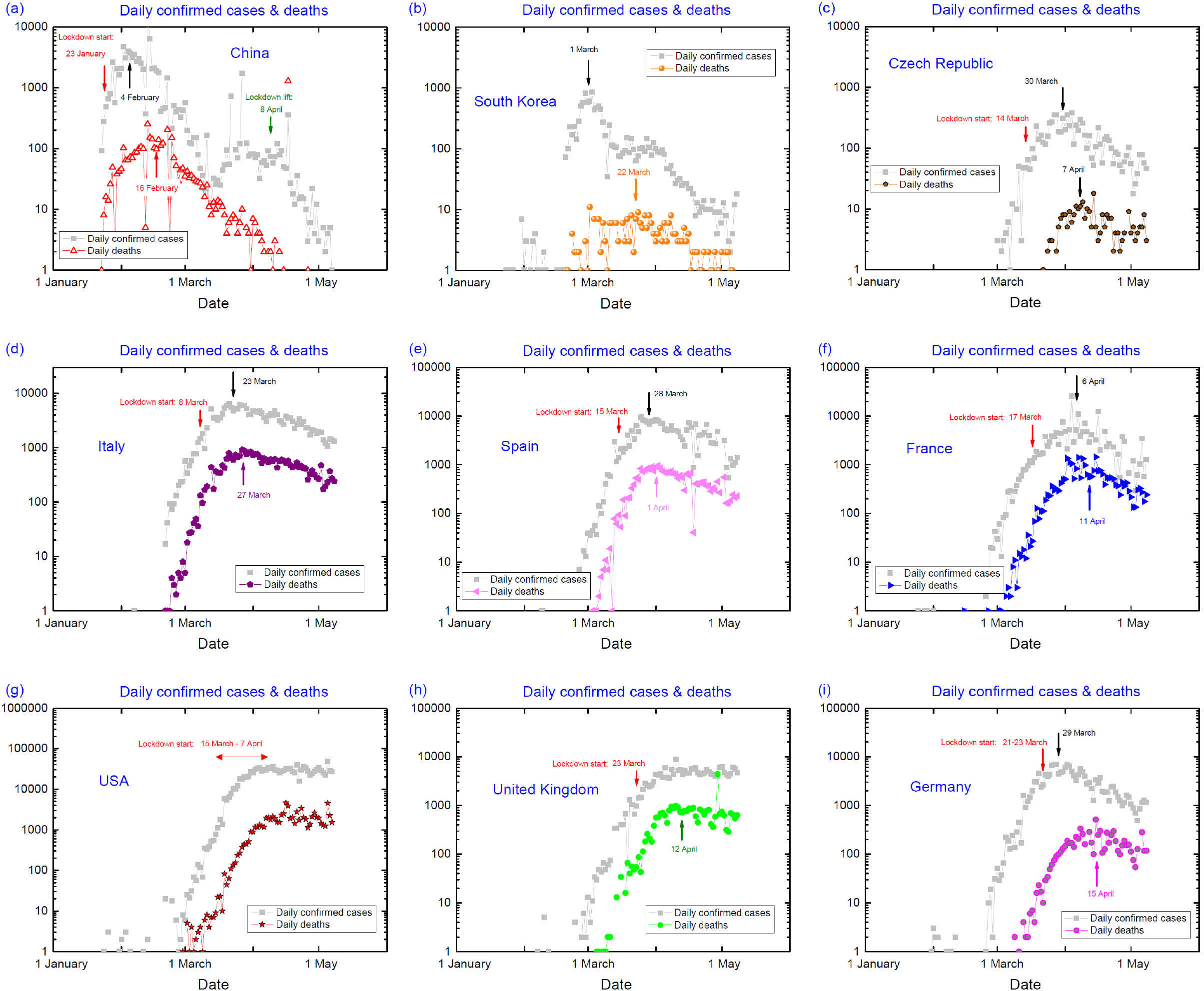
*Time variation of daily confirmed cases and deaths in a selection of nine countries [26],[27],[28],[29],[30],[31],[32]*

In Czech Republic, Italy, Spain, France, and Germany the epidemic peak has been observed for both numbers of daily cumulative cases and deaths. However, in Italy a few weeks after the peak the number of daily deaths decreased with a slower rate than in China. In the United States of America and in the United Kingdom, both daily confirmed cases and death tolls have been saturating in the last weeks of April, and no clear peak emerged so far from the plateaus. In all of these western countries (except Czech Republic), daily confirmed cases and death numbers are still several orders of magnitude higher than those in China when lockdown was lifted, or than those in South Korea after the epidemic peak.

Figure 3 focuses on a comparison between two neighbors from the Scandinavian Peninsula: Sweden and Norway. Beyond their geographic and climatic similarities, both countries are quite comparable in term of population and density (10 million inhabitants and 450 000 km^2^ for Sweden, 5.4 million inhabitants and 385 000 km^2^ for Norway [35]). The pandemic started in these two territories almost simultaneously, as shown by the sudden increase of daily confirmed cases after the 1 March [Figure 3(a)] and of the daily deaths [Figure 3(b)] two weeks later in both countries. Two opposite strategies were followed by Sweden and Norway to face the pandemic. Sweden was guided by the target to let the pandemic spreading over the territory, so that herd immunity [4],[5],[6] is achieved in a ‘natural’ manner [36]. A few days after the first reported cases, Norway applied strong measures and lockdown was set on 12 March, i.e., before the first reported death [33]. The effects of these measures are visible in Figure 3, with broad maxima centered on 27 March in the daily cumulative cases, and on 10 April in the daily deaths. In the beginning of May, the first epidemic wave has almost ended in Norway, with < 1 ‘average’ daily deaths and < 30 daily confirmed cases. In Sweden, without lockdown the number of daily reported cases and deaths increased before reaching a plateau with 300-900 daily confirmed cases and 10-200 daily deaths. A large noise in the data indicates a difficulty to collect data in this country. In the beginning of May 2020, ≈ 200 cumulative deaths were reported in Norway and > 3 000 cumulative deaths were reported in Sweden, where the first epidemic wave was about to continue and to lead to a higher death toll. The comparison between Sweden and Norway is a direct illustration of the human cost of the herd immunity strategy.

**Figure 3:**
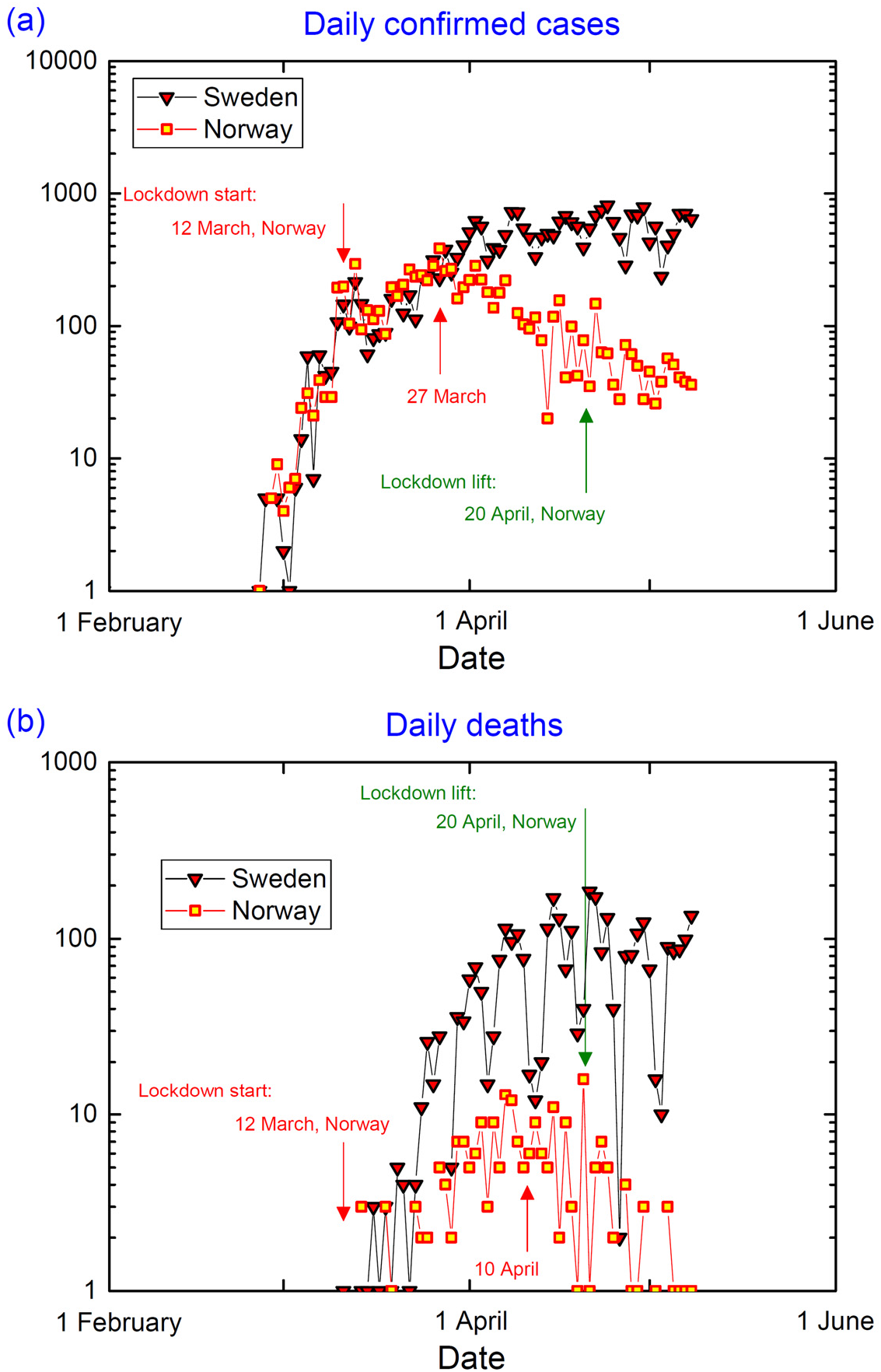
*Comparison of daily confirmed cases and deaths in Sweden and Norway [33],[34]*.

### 3.2- Phenomenological description

#### 3.2.1- Definition of three epidemic regimes

Figure 4 focuses on the variation of cumulated death and daily death tolls normalized per 100 000 inhabitants for four countries: China, South Korea, Italy and France. In the non-normalized graphs plotted in Figure 1(c,f,i), the date corresponding to the day *D* = 0 of the ‘shifted’ time scale was defined by adjusting the extrapolation of the exponential-growth regime to *N* = 1 cumulative death. Since the criterion *N* = 1 is not proportional to the population, if we compare two countries of different populations and hit at the same time by the pandemic, a later ‘shifted’ date *D* is artificially defined for the country of smaller population. The consideration of data normalized with regard to the population permits to avoid this artefact. In Figure 4, but also in the next graphs presenting tolls normalized per 100 000 inhabitants, the day *d* = 0 is defined as the extrapolation of the exponential-growth regime to *n* = 0.001 cumulative deaths / 100 000 inhabitants. Normalized death tolls will be systematically considered in Section 3, where a quantitative comparison of the pandemic spread is proposed for thirty-two countries worldwide.

**Figure 4:**
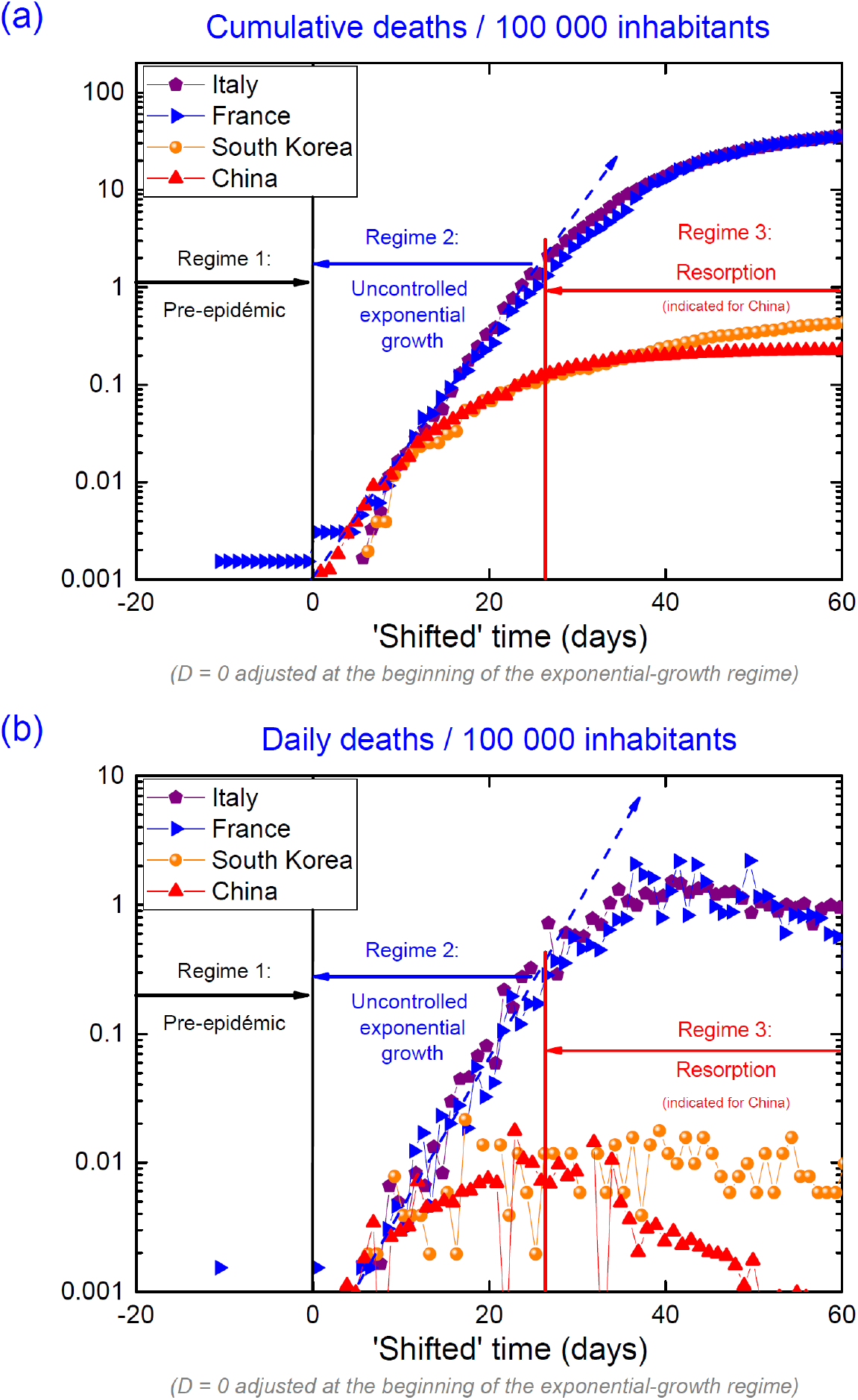
*Identification of three regimes in a single-wave epidemic propagation*.

Figure 4 indicates that three regimes can be identified in a single-wave propagation of the pandemic:

- Regime 1: pre-epidemic phase, where zero or a few isolated cases are reported, and where the propagation rhythm is zero or weak. In this regime, the epidemic is kept at bay.
- Regime 2: exponential- and uncontrolled-growth phase. The propagation is wild and not slowed down, the numbers of reported cases and deaths are increasing exponentially, and a universal law is followed. In the semi-logarithmic-scale graphs presented here, all countries show a linear variation of similar slope when they are in regime 2. Both cumulative and daily case numbers increase exponentially in this regime.
- Regime 3: resorption phase. After a deviation from the exponential-growth regime 2, this phase corresponds to a decay of the epidemic propagation. Here, we define the transition (in fact a broad crossover) from regime 2 to regime 3 at the date when the numbers of daily death tolls passes through a maximum, which is identified as the epidemic peak. The decline of the epidemic propagation ends asymptotically by a saturation of the cumulative death number. Regime 3 can be the result of several causes: i) the success of a national policy in the slowing down of the virus propagation
- (mitigation, containment, lock-down, vaccination etc.), ii) a number of contaminated cases approaching the population number (if there is fewer persons to contaminate, there will be less infected people), meaning that a collective immunization is approaching, iii) particular local conditions (climate, hot temperatures) unfavorable to the virus, or iv) a failure (voluntary or not) in the counting system.

While the transition between regimes 1 and 2 is sharp and fast, that between regimes 2 and 3 is progressive and spreads over several weeks.

#### 3.2.2- Two-parameters description of first-epidemic waves

A universal behavior is observed in regime 2 where the epidemic dynamics is out of control. An open question is whether the transition between the regimes 2 and 3 is also universal, or if it depends on local specificities, as an interaction rate in the population, the efficiency of social distancing, mitigation, containment or lockdown when applied. Here, extrapolations are made with the crude, but perhaps not unrealistic, assumption that for each epidemic wave the transition between regimes 2 and 3, and then the resorption in regime 3, are similar to those reported for the first epidemic wave in China. The evolution of the cumulative death toll is estimated using a smoothed curve constructed from the Chinese cumulative death variation.

For each wave, two parameters are adjusted: an offset in time and a scaling factor *F* in the death number. The scaling factor *F* corresponds to the ratio of cumulative deaths at the beginning of the lockdown and at the end of the epidemic wave. Efficient lockdown measures are associated with a smaller value of *F*. Graphically, in a semi-logarithmic scale this simply corresponds to a translation of the dashed black line initially adjusted on China’s data. These fits summarized in Figure 5 do not intend to precisely predict the final number of cumulative deaths for an on-going epidemic wave. They show that the dynamics of a COVID-19 wave is similar in the different countries, and they indicate the typical time scales and the orders of magnitude of the final death tolls expected at the end of an epidemic wave.

**Figure 5:**
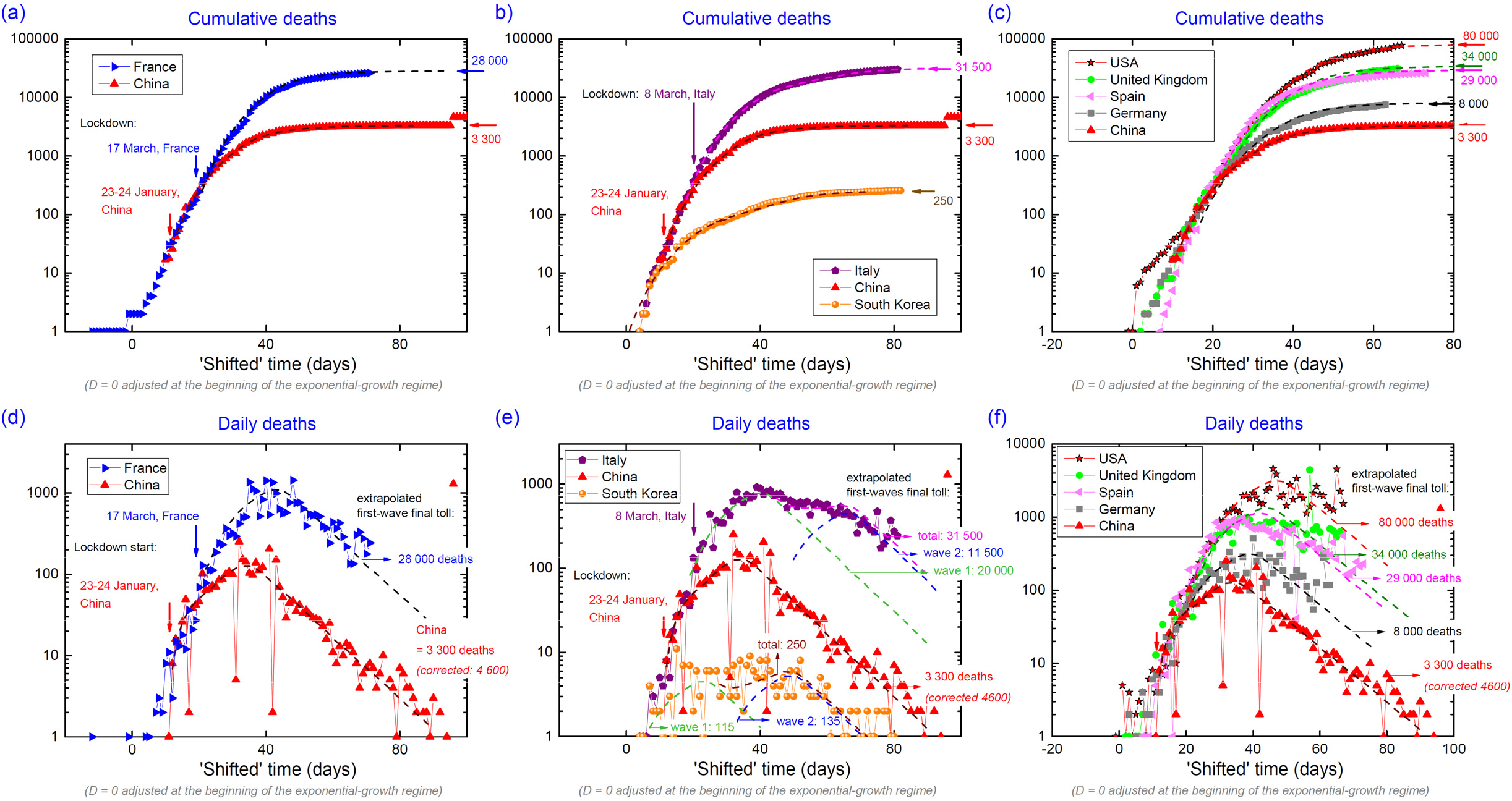
*Extrapolation of (a) cumulative and (d) daily death numbers in France, assuming a unique wave similar to that reported in China. Extrapolation of (b) cumulative and (e) daily death numbers in Italy and South Korea, assuming two successive epidemic waves, each one being similar that reported in China. Extrapolation of (c) cumulative and (f) daily death numbers in the United States of America, the United Kingdom, Spain and Germany, assuming a unique wave similar to that reported in China*.

For most of the considered countries, at this stage (data up to 8 May 2020) the assumption of a single epidemic wave is sufficient to fit the data within first approximation. Higher final numbers than those given here will probably be observed, due to resurging waves or late death-toll corrections. For France, the best fit to the data, with a factor *F* = 160, corresponds to a final number of 28 000 cumulative deaths for the first wave. Similarly, fits are compatible with extrapolations to 8 000 final cumulative deaths in Germany, 29 000 final cumulative deaths in Spain, 34 000 final cumulative deaths in the United Kingdom, and 80 000 final cumulative deaths in the United States of America. For Italy and South Korea two successive waves are used to describe an anomalously-long epidemic plateau. For Italy, the fit is compatible with a first wave of 20 000 final cumulative deaths, with a maximum peaked 40 days after the start of regime 2, and a second wave of 11 500 final cumulative deaths, with a maximum peaked 60 days after the start of regime 2. For South Korea, the fit is compatible with a first wave of 115 final cumulative deaths, with a maximum peaked 20 days after the start of regime 2, and a second wave of 135 final cumulative deaths, with a maximum peaked 50 days after the start of regime 2. The results from these phenomenological fits are in good agreement with those from more sophisticated models (see for instance [15]).

Figure 6 summarizes the fits made here for the United States of America, the United Kingdom, Spain, Italy, France, and Germany. In this graph, the daily death tolls are plotted as function of non-shifted date. Two months after an epidemic peak of 100-200 daily deaths, China ended the lockdown in Wuhan, where the virus has been the most active, on 8 April. At this date, the daily death number was of the order of 1. An almost-constant negative slope in the evolution of the daily death numbers, in this semi-logarithmic plot, was reported during the two months after the epidemic peak in China. It indicates an exponential decay of the daily death number with time. Assuming a similar decay for the other countries, longer lockdown duration is expected in countries where the epidemic peak reached a higher level. However, contrary to the Chinese strategy, in beginning of May 2020 most of the Western countries were planning to lift their lockdown soon after the epidemic peak, at a date where hundreds of daily deaths were still reported.

**Figure 6:**
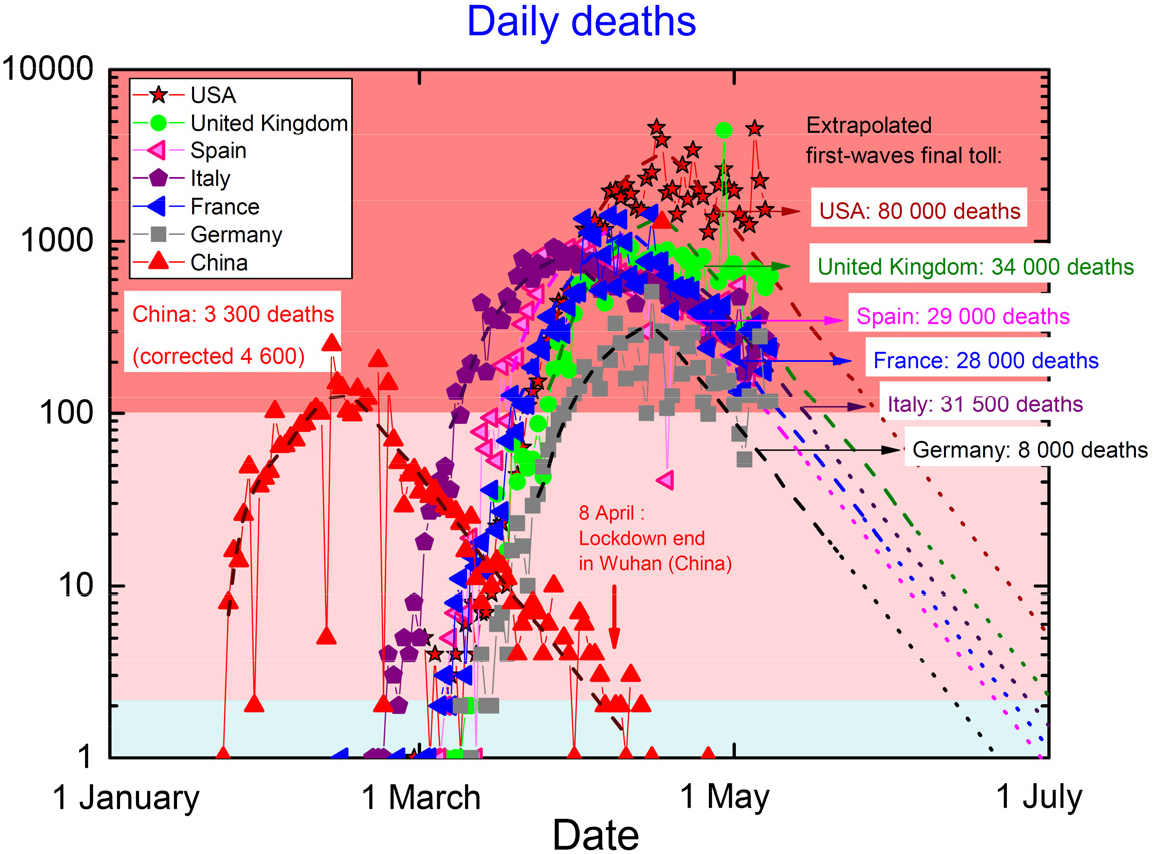
*Fit to the daily death tolls as function of date in the United States of America, the United Kingdom, Spain France, Italy and Germany, assuming epidemic waves dynamics similar to that reported in China*.

### 3.3- Comparison of the propagation of the pandemics for a selection of countries

While the raw data presented in Section 1 spotlight the countries with a large population, in this Section we consider graphs where the cumulative and daily death tolls have been normalized with regard to the population [35] (see Section 2.1). This permits to quantitatively compare the spread of the pandemic in countries of different populations.

Figure 7 presents the cumulative and daily death tolls normalized per 100 000 inhabitants as function of a ‘shifted’ time, for a selection of thirty-two countries worldwide (data ending in the beginning of May 2020). Complementary plots of confirmed cases and deaths data for these countries are presented in the Supplementary Materials (Figure S1). In most of the countries, a similar exponential-growth regime is observed in the time-evolution of the death numbers. The efficiency of the measures taken in Asia is revealed spectacularly in the graphs of Figure 7: the death tolls per 100 000 inhabitants are two orders of magnitude smaller in Asian countries than in the western countries listed above. A maximum of 3-4 daily deaths / 100 000 inhabitants has been reported in Belgium, which is the mostly-affected country. Maxima of 1-2 daily deaths / 100 000 inhabitants were reported in several Western Europe countries, as Spain, France, Italy, United Kingdom and Sweden, and in the United States of America. The spread of the pandemic is heterogeneous in Europe, and countries as Greece, Czech Republic, and Norway succeeded to contain it to < 0.2 daily deaths / 100 000 inhabitants, which is a few times higher than the rates < 0.05 daily deaths / 100 000 inhabitants reported in Asian countries. In the Supplementary Materials (Section S4), we show that similar inhomogeneity can be observed at a national scale, once regions and departments are considered separately. In other parts of the world (Africa, South America, Australia), the reported death numbers indicate a situation in-between that in Asia and that in mostly-affected western countries, due to the combination of late arrivals of the virus on the territory and possible local specificities (density of population, weather, etc.).

**Figure 7:**
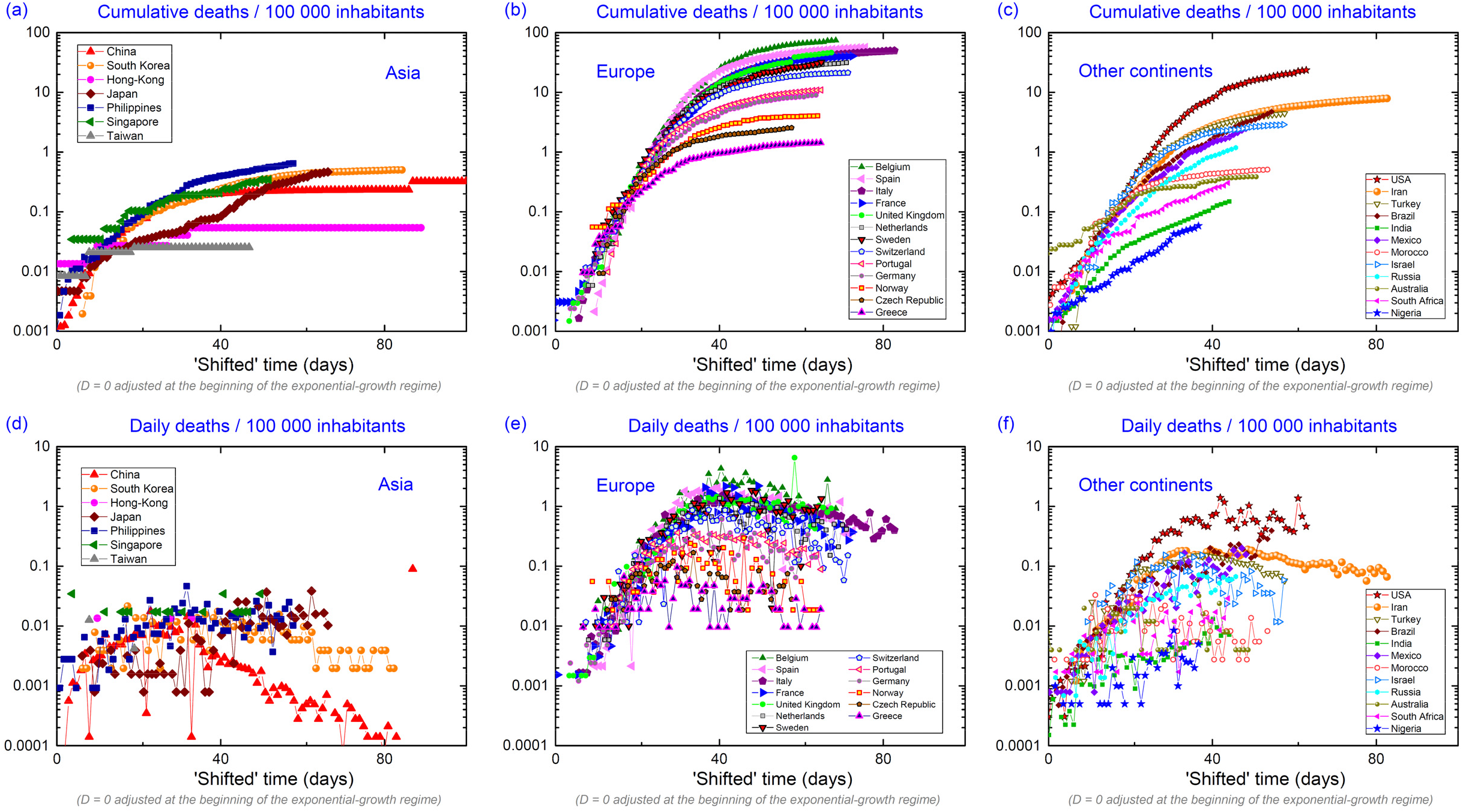
*Cumulative and daily death tolls per 100 000 inhabitants for a selection of countries*.

The world map in Figure 8 indicates the delays in the worldwide propagation of the COVID-19. The ‘shifts’ in time used in the data plotted in Figure 7, in relation with the delayed onset of the exponential-growth regime 2 [see Figure 4], are indicated for the countries considered here. This Figure shows that, four months after the first cases reported in China, all parts of the globe have been hit by the pandemic. After Asia, the pandemic arrived in South-West Europe and then expanded to the North and East of Europe, to the United States of America, and finally to the rest of the world.

**Figure 8:**
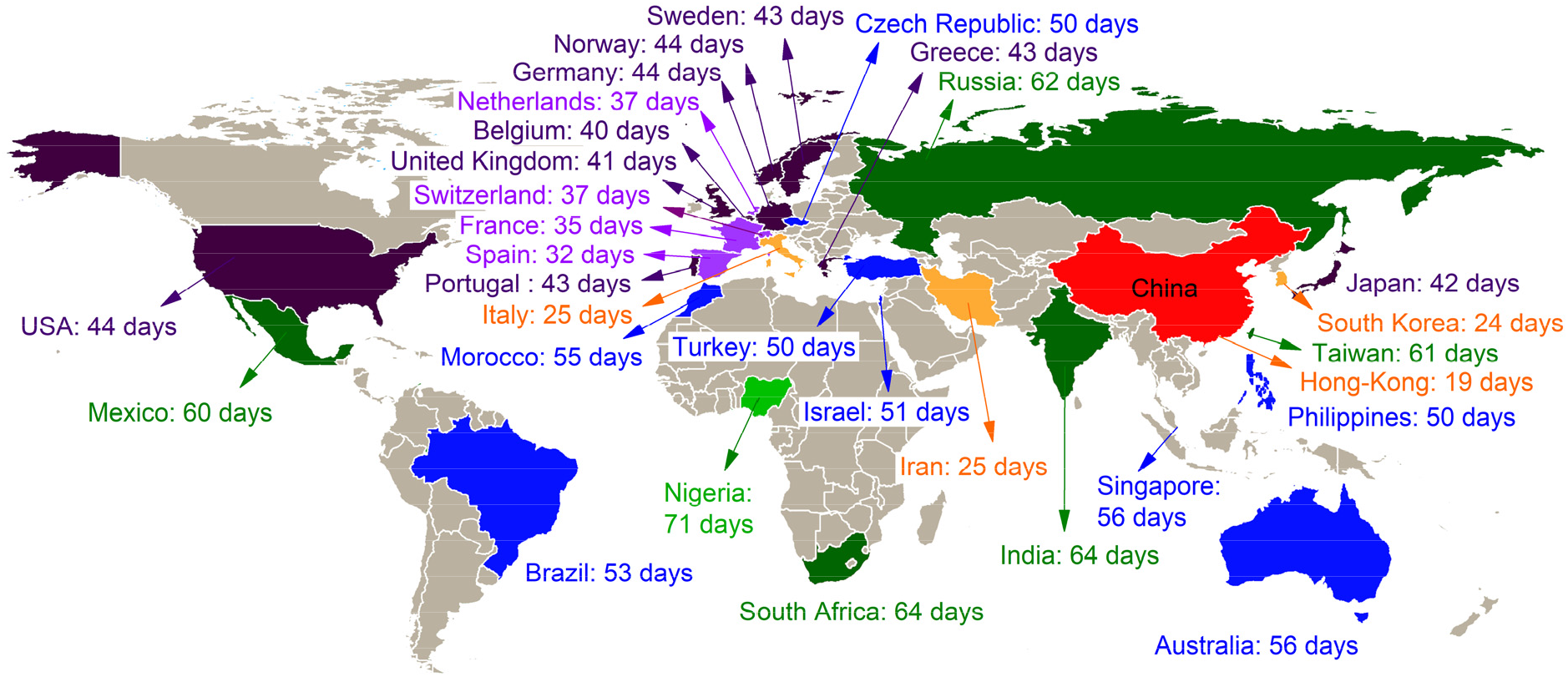
*World map and shifts in days of the beginning of the epidemic exponential-growth regime 2, for a selection of thirty-one countries, in comparison with China*.

Table 1 summarizes the situation for the countries considered here. It shows that the pandemic spread in the different countries is weakly-correlated with the date of arrival of the pandemic. This means that the experience gained by early-hit countries did not benefit to all lately-hit countries. The countries considered here are classified in a four-group scheme, depending on the degree of spread of the pandemic in their population on 8 May 2020:

- *<u>Group A:</u>* Taiwan, Hong-Kong, Japan, China, Singapore, Philippines, South Korea, Nigeria, South Africa, India, India, Australia, Morocco. The pandemic was contained to low levels, with less than 0.5 cumulative death / 100 000 inhabitants. In most of these countries measures were taken in reactive and efficient way. In some of them, a late arrival of the virus combined with local specificities (weather, age of population, etc.) perhaps helped to keep low death tolls in the beginning of May 2020.
- *<u>Group B:</u>* Russia, Mexico, Greece, Brazil, Czech Republic, Israel, Turkey, Norway. These countries have been ‘weakly’ affected by the pandemic, with between 0.5 and 1 cumulative death / 100 000 inhabitants. In Europe, the results obtained in Greece, Czech Republic, and Norway contrast with those from their neighbors, most of them being in group D. In the beginning of May 2020, the daily death tolls in Brazil, Mexico, Russia continue increasing, and these countries may later downshift to Group C.
- *<u>Group C:</u>* Iran, Germany, Portugal. From the official tolls, these countries are in a better situation than the countries from group D. However, the situation is not optimal, since between 7 and 9 cumulative deaths / 100 000 inhabitants were reported, which is more than a factor 10 higher than in the countries from group A.
- *<u>Group D:</u>* United States of America, Switzerland, the Netherlands, Sweden, United Kingdom, France, Italy, Spain, Belgium. This ‘group is composed of countries from Western Europe and North America. Fashionable theories (herd immunity scenario [4],[5],[6]) and a confidence in health system perhaps led to a delayed state reaction against the pandemic propagation. Lockdown measures were applied late, when the numbers of cumulative deaths were already high, leading to much higher epidemic peaks and cumulative death tolls than in the countries from the groups A-C. A maximum of 3-5 daily deaths / 100 000 inhabitants was reported in Belgium, and maxima of 1-2 daily deaths / 100 000 inhabitants were reported in Spain, France, Italy, Sweden, United Kingdom and United States of America. In the beginning of May 2020, Sweden was the last strongly-affected country having the strategy to reach herd immunity without lockdown measures. Its situation may continue worsening till herd immunity is achieved or till the Swedish government changes its strategy.

**Table 1:**
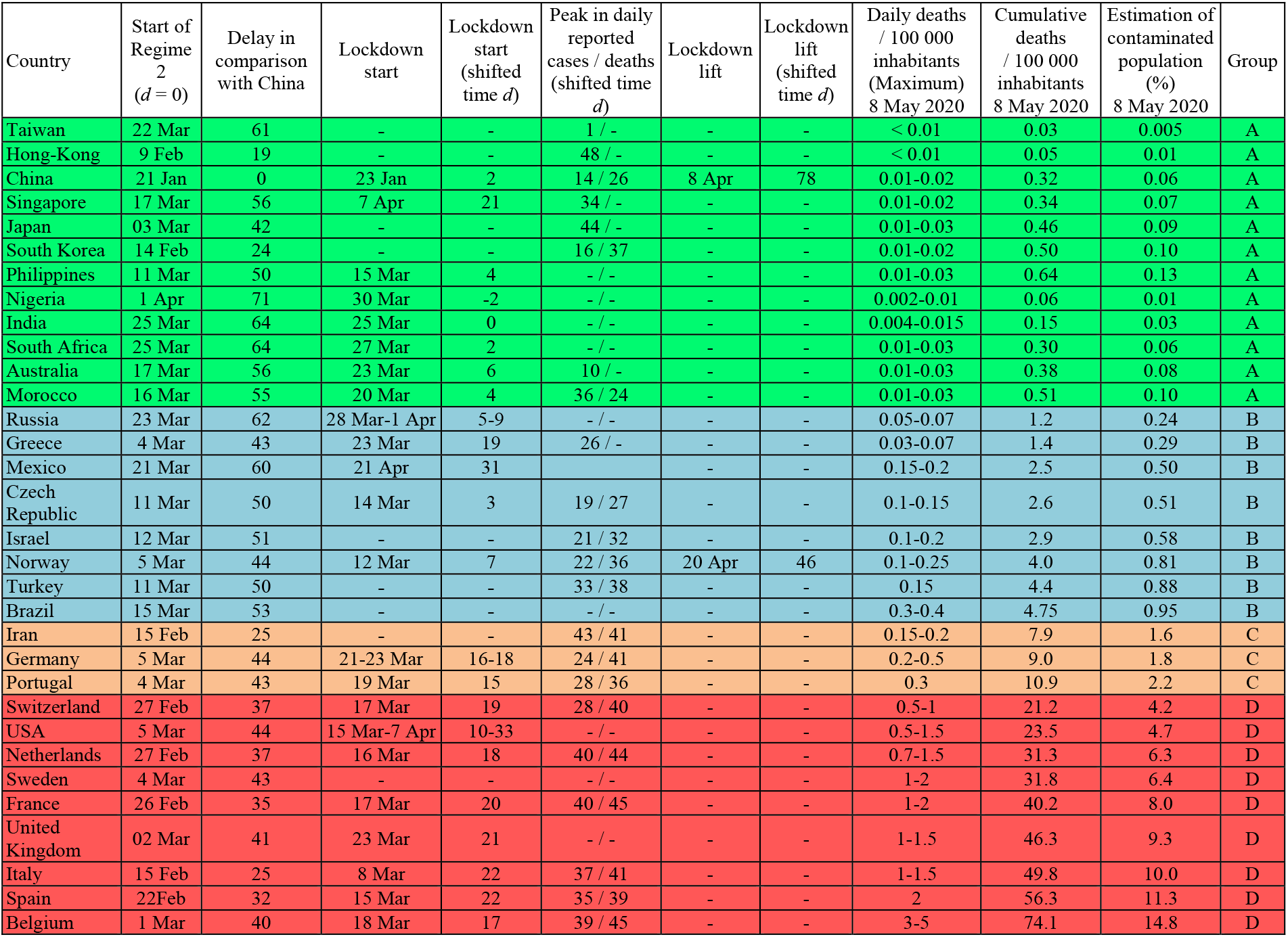
*Comparison of characteristic times in the propagation of the pandemic, deaths rates (maximum daily deaths and cumulative death tolls, normalized per 100 000 inhabitants, observed till the 8 May 2020) for the selection of thirty-two countries considered here. Lockdown start and lift dates are from [26], [27], [28], [29], [30], [31], [32], [33], [37], [38], [39], [40], [41], [42], [43], [44], [45], [46], [47], [48], [49], [50], [51]. Assuming a fatality rate of 0.5 %, the percentage of population already hit by the virus is estimated for each country. The countries are ranked into four groups A-D, depending on the degree of propagation of the pandemic in their population*.

Table 1 also indicates that a clear relation exists between the earliness of application of lockdown measures and the efficiency to contain the pandemic spread. From most of the countries considered here, the epidemic peak, i.e., the center of the maximal plateau in the daily deaths, variation is observed 20-25 days after the application of lockdown. The effects of a lockdown are, thus, observable quite late, which indicates the importance of applying it immediately after the start of an epidemic wave. The Section S2 in the Supplementary

Materials details the dramatic effects of a delay in the application of a lockdown, once an exponential-growth regime 2 is established. Early lockdown dates, before or a few days after the *d* = 0 start of the exponential-growth regime 2, characterize the countries from Group A (those who applied lockdown). On the contrary, all countries from Group D (with the exception of Sweden) applied a late lockdown, about three weeks after the start of the exponential-growth regime 2. Most countries from Groups B and C are in an intermediate situation.

Figure 9 presents the time variation of the ratio between the cumulative deaths and confirmed cases for the selection of countries considered here. The ‘shifted’ times used in this graph were defined by considering normalized death tolls per 100 000 inhabitants (see Figure 7). Even in the case of a perfect ‘measurement’, where the cumulated deaths and confirmed cases would be well-estimated, their ratio would not be constant with time, due to the time delay between contaminations and deaths. For a perfect ‘measurement’, this ratio would lead asymptotically to the fatality rate of the epidemic at the end of an epidemic wave. However, this rate is not universal, since it can vary from one country to another, due to different weather conditions, population characteristics (age, obesity, density, etc.), medical care means, and possibly virus mutations. In real life, measurements are imperfect and the means to detect COVID-19 cases are more or less efficient, depending on the country. The large scattering of data in Figure 9 mainly results from these counting limitations. Since cumulative death tolls are expected to be more reliable than confirmed case tolls, the ratio at the end of an epidemic wave reaches a value higher than the fatality rate when measurements are imperfect. We can suspect that a small country doing a high number of tests may be able to reach a ratio close to the fatality rate. This may be the case of Hong-Kong, for which the ratio converges to 0.4 % at the end of the epidemic wave, which may be compatible with a fatality rate of ≈ 0.5 %. This value corresponds to estimates of fatality rates, ranging from 0.5 to 1.5 %, proposed in Refs. [9],[10],[52]. We note that a smaller ratio 0.1 % was observed in Singapore in the beginning of May 2020, but this value may increase since the epidemic wave is still on-going in this country.

**Figure 9:**
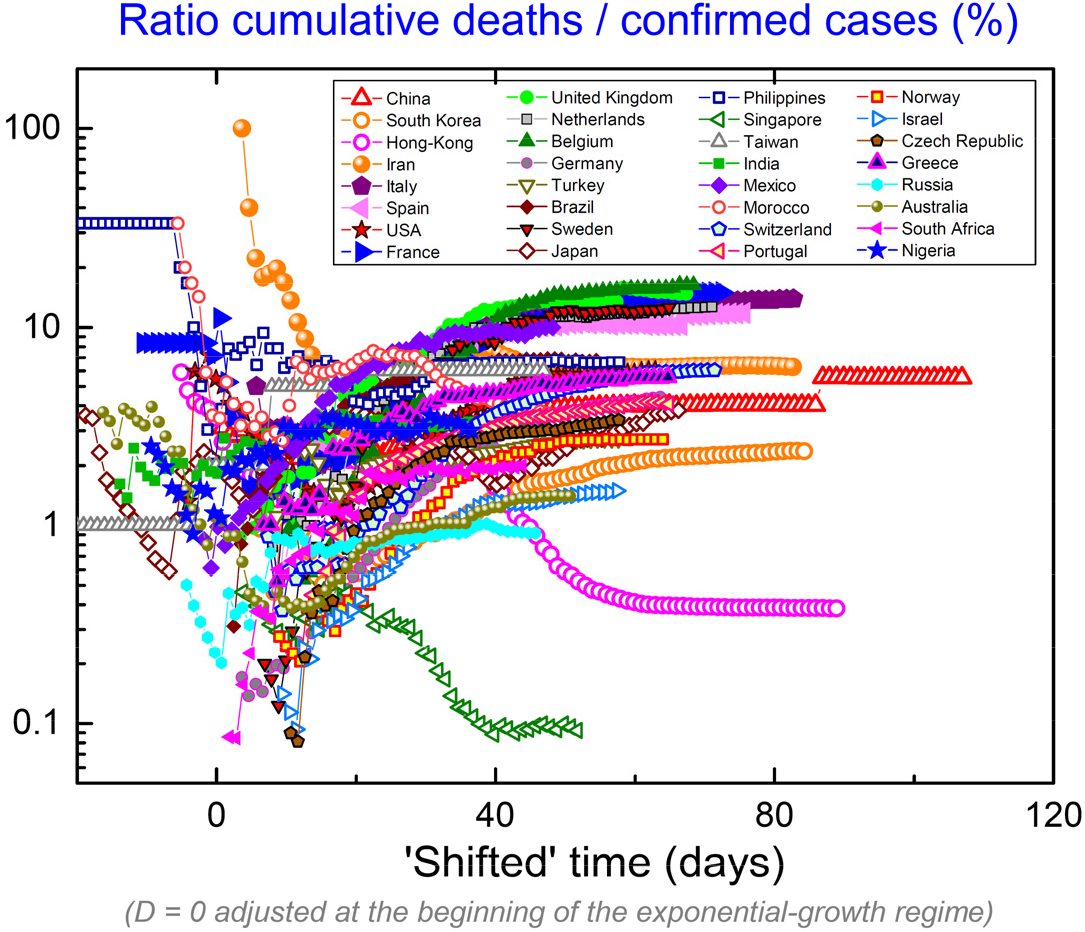
*Ratio between cumulative deaths and confirmed case tolls for the selection of countries considered here*.

From this rough, but presumably reasonable, assumption of a fatality rate of 0.5 %, we can estimate the order of magnitude of the already-infected part of the population in each country by applying a factor 200 to the cumulative death number. Due to the delay between contaminations and their consequences (including deaths), this estimation is more appropriate for countries at the end of an epidemic wave. Table 1 shows the estimated proportions of the population infected by the coronavirus SARS-CoV-2 at the date of 29 April 2020: less than 0.1 % in countries of the group A, between 0.1 and 1 % in countries of the group B, between 1 and 2 % in countries of the group C, and more than 4 % in countries of the group D. A maximum of 15 % of infected people is estimated for Belgium, which is the mostly-affected country. For all countries from Group D, these tolls remain far below from the proportion of 60 % expected to achieve herd immunity. In the mostly-affected parts in Europe (as department Bas-Rhin in France, see Section S4 in the Supplementary Materials), a maximum of 100 cumulative deaths per 100 000 inhabitants was reported and we can estimate that 20 % of the population was contaminated. In these highly-affected areas, the number of contaminated cases is still far from the proportion of 60 %. On 27 April, New-York City was one of the mostly-affected areas in the world, with 19 561 cumulative deaths reported [24], which corresponds to 0.235 % of its population of 8.3 million inhabitants [53]. Assuming a fatality rate of 0.5 %, we can estimate that 47 % of the population was infected by the virus. New-York city may be soon the first area with several million inhabitants where herd immunity is achieved. These rough estimations also confirm that herd immunity would be an option of very high human cost if achieved at a worldwide scale (see also Section S3 in the Supplementary Materials).

## 4- Discussion

The Asian countries from the group A succeeded to contain the spread of the pandemic to low levels. In China, lockdown was applied early after the identification of a starting epidemic wave. An epidemic peak with a rate of 100-200 deaths per day was reached 20 days later, and lockdown was lifted 80 days after its application, while an average rate of < 1 death per day was reported. In South Korea, an early reaction permitted to isolate most of the contagious cases and to early break the pandemic dynamics, with a total of 250 cumulative deaths counted at the beginning of May 2020. To keep a low number of contagious cases, and thus, a low number of new contaminated cases, the strategy of Asian countries of Group A can be summarized by the main measures [54]:

- systematic tests of the population combined with a fast isolation of new contaminated cases
- a massive use of protection masks
- a strict surveillance of national frontiers, with quarantine imposed to all new incomers. The results obtained so far can be considered as a validation of this strategy. They may allow continuing an economic activity without risking the resurgence of large epidemic waves. Bilateral agreements between ‘safe’ countries may permit to reopen progressively the frontiers and to restart economic exchanges. For these reasons, but also since risky strategies may be followed in Western countries (see below), Asian countries are natural candidates to be the ‘winners’ of the worldwide economic crisis starting in 2020.

In the countries from Group B, after the end of the first epidemic wave, the levels of contagious people are low enough to hope avoiding a second devastating wave, once appropriate measures are taken. A similar method to that applied in Asian countries may constitute a healthy strategy for the forthcoming weeks/months. A difficulty will be to avoid contaminations imported by their neighbors from Groups C and D.

In the countries from Groups C and D, euphoria subsequent to the decay of reported cases after the first epidemic peak could lead to a worsening of the situation. A patient approach, with a lockdown lift in end of June, when the number of daily death tolls would be of the order of a few units per country (see Figure 6), may allow to safely reach a situation similar to that of China and South Korea after their first epidemic waves. On the contrary, a hasty-lockdown-lift strategy, i.e., the end of lockdown measures while the number of contagious people would remain high, may lead to a grand second wave. Stopping a lockdown when the epidemic is still active constitutes a risky strategy, which could void in a few days the results obtained by a one- or two-months-long lockdown. A re-opening of the frontiers inside the Schengen area and a non-massive use of protection masks may constitute additional difficulties to avoid large resurgent epidemic waves in the Western Europe countries from Group D.

The graphs and the phenomenological descriptions presented here emphasize the importance of applying reactive and efficient measures against the propagation of the COVID-19 pandemic. Such intensive effort may be needed as long as herd immunity is not achieved, either by a global vaccination campaign or by a free (voluntarily or not) spread of the pandemic in a population. An application of the methods which proved to be successful in Asia, rather than the tentative of alternative and risky methods, is suggested for the countries facing strong epidemic waves. Additional complications could come from a seasonality of the virus, which would prevent reaching herd immunity without vaccine.

In beginning of May 2020, several questions are still open:

a. Are the measures taken in Asian countries, as China and South Korea, sufficient to limit epidemic resurgences to ripples associated with a ‘few’ tens or hundreds of cumulative deaths? Is it possible to maintain the pandemic to such low level during 10-20 months, i.e., the expected timescale for a vaccine available in large quantities?
b. How will Western Europe countries proceed to try avoiding new devastating waves? Will they apply similar measures than the Asian countries, or will they experiment alternative methods? In particular, can a hasty lift of lockdown lead to the resurgence of epidemic waves of significant magnitude? In such case, could held immunity be achieved, and would the associated sacrifice of hundred thousand lives in each country sufficient for a fast economic reboot?
c. How the situation will evolve in the countries of South America, Africa and Oceania, where the late arrival of the pandemic, possibly combined to local specificities (weather, age of population, etc.), permitted them not to be heavily-hit in the beginning of May 2020?

## 5- Conflict of Interest

No conflict of interest.

## Data Availability

data available on reasonable request to authors

## Supplementary Materials

### S1- Supplementary graphs

**Figure S1:**
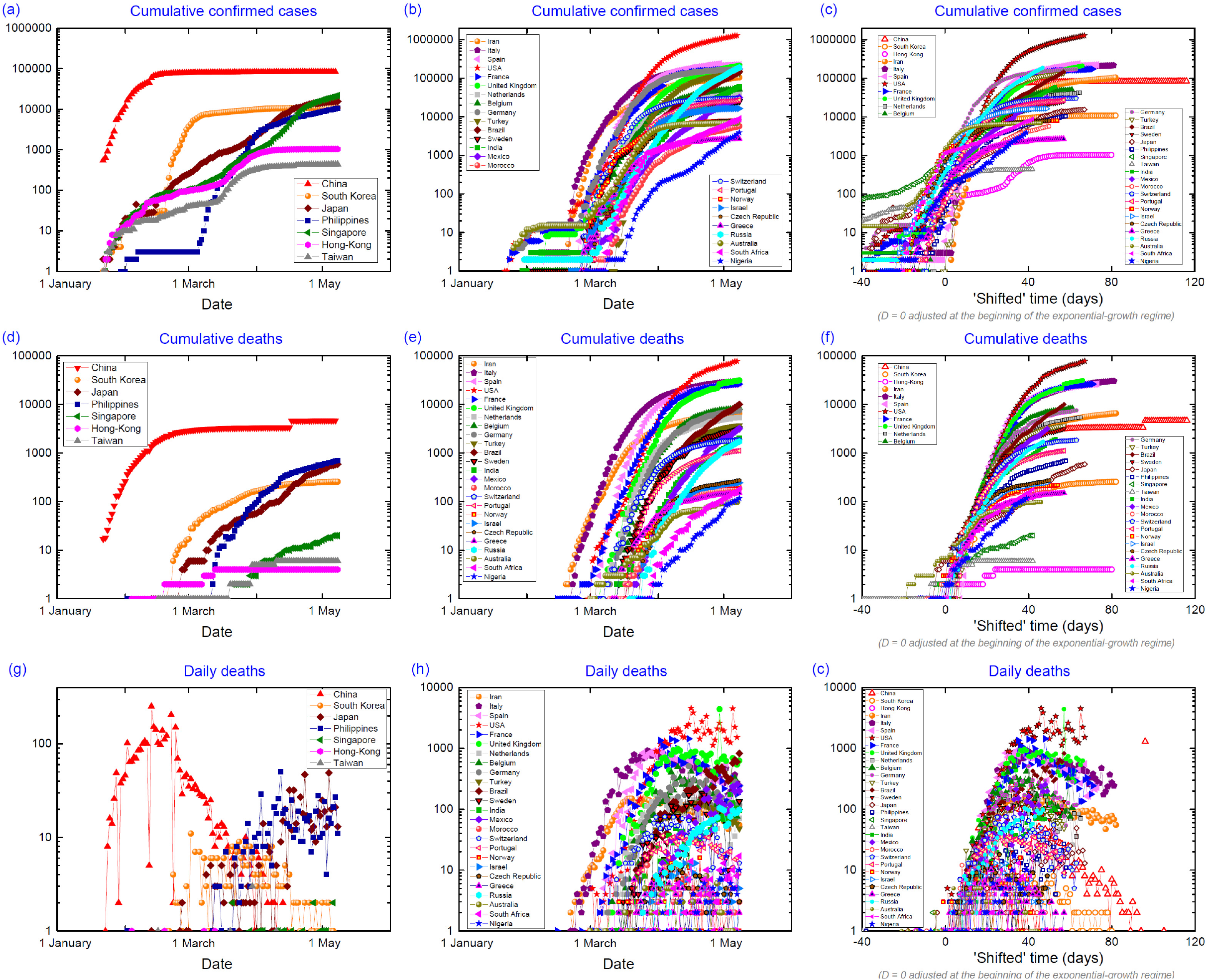
*Comparison of the propagation of the pandemic in a selection of thirty-two countries*.

**Figure S2:**
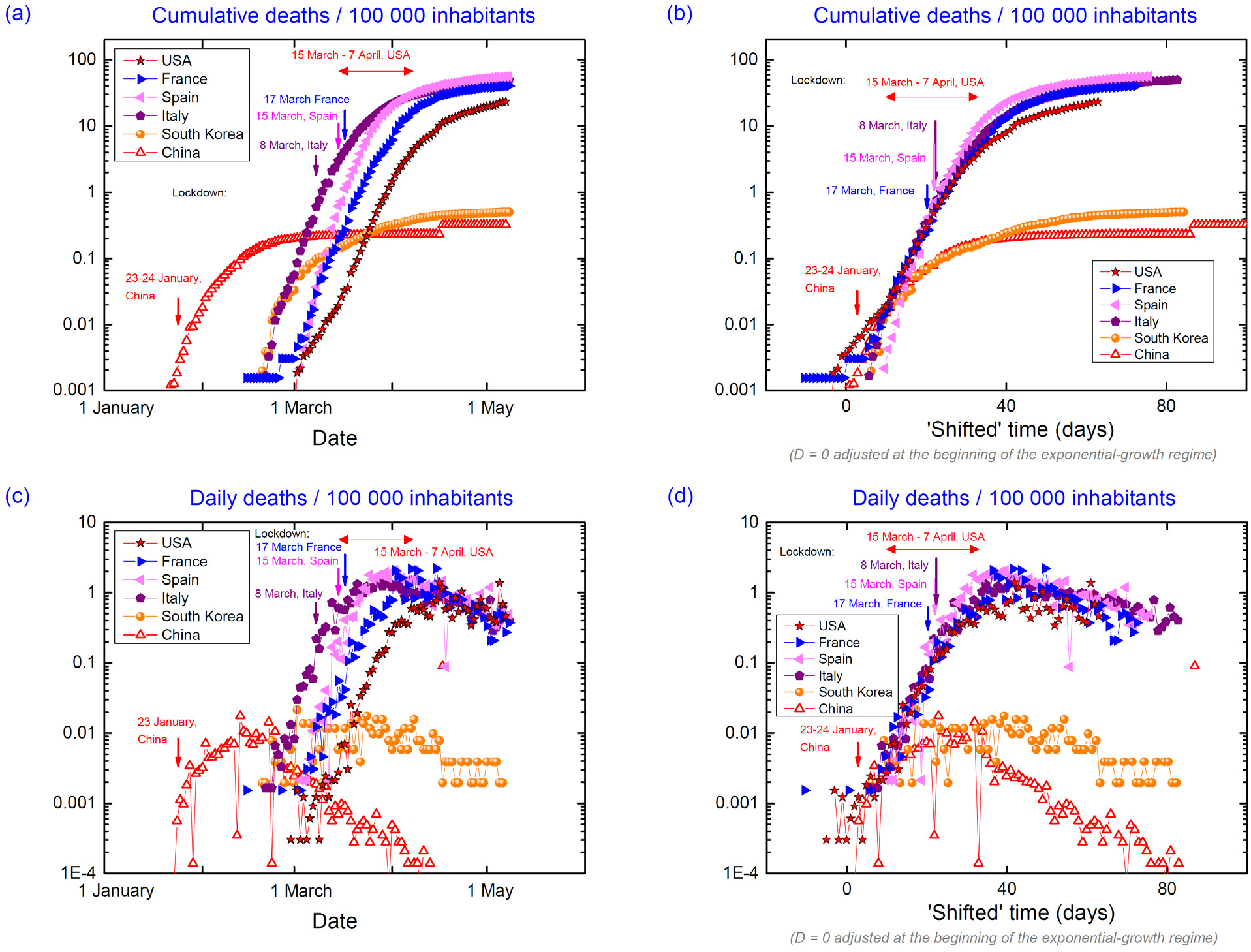
*Evolution of cumulative and daily death tolls per 100 000 inhabitants in China, South Korea, Italy, Spain, France (mainland) and the United States of America*.

### S2 - Consequences of a delay in the lockdown start

Figure S3 shows the effects of a delay in the establishment of lockdown on the final cumulative death toll at the end of an epidemic wave. Figure S3(a) shows that the increase of the cumulative deaths in a country in the exponential-growth regime 2 (fit done to the data in Italy) follows the exponential law:

*N* = exp(0.2924*D),

where *N* is the cumulative death number and *D* the day in the ‘shifted’ time scale defined in Section 1.

Figure S3(b) presents an extrapolation of the final cumulative death number in France. It was done assuming two hypotheses:

**Figure S3:**
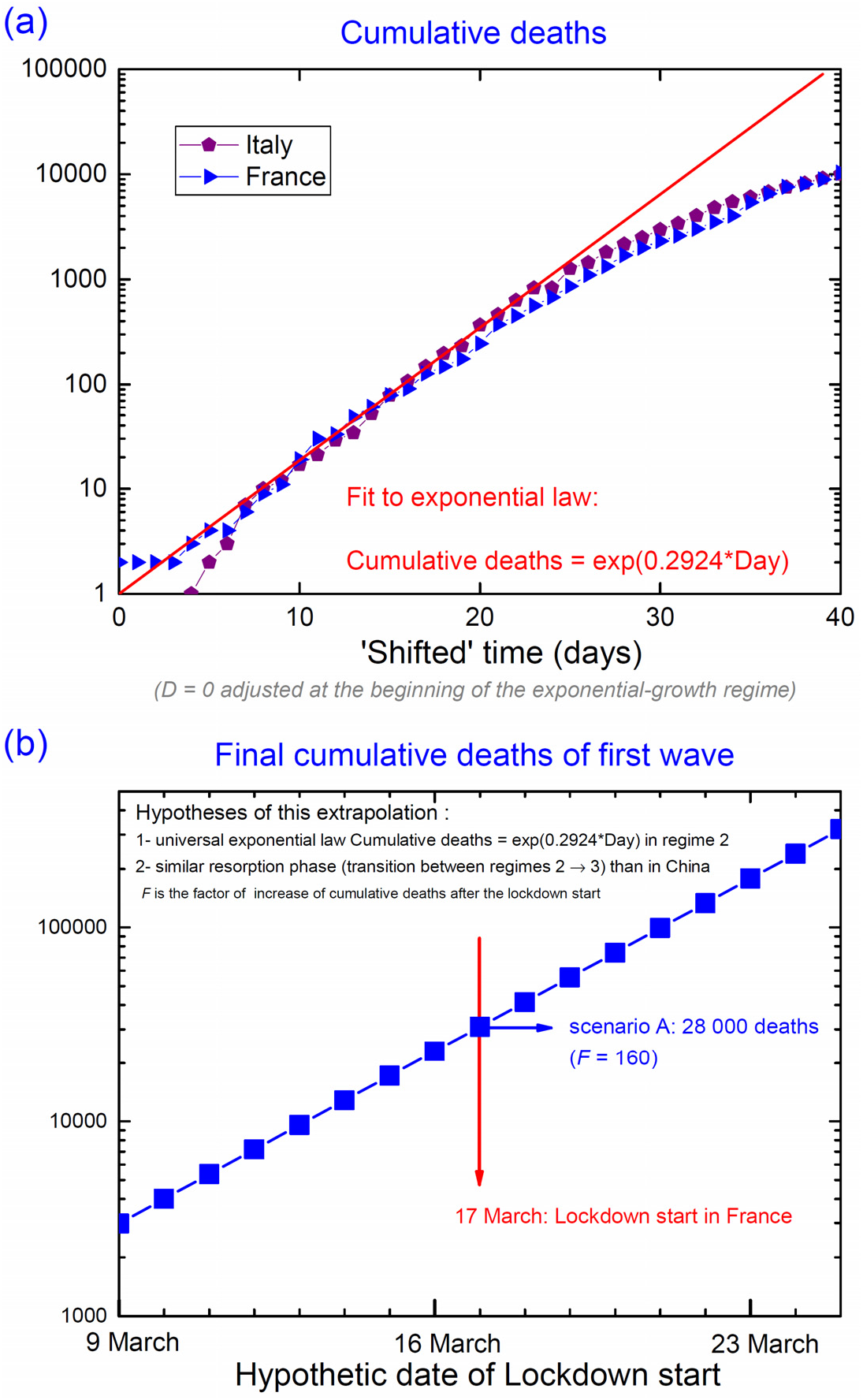
*(a) Fit by an exponential law of the cumulative deaths variation in regime 2. This universal law is followed by all countries in this regime*. *(b) Extrapolation of the final cumulative death number expected in France, as a function of the starting date of lockdown. A similar epidemic decline as that reported in China, and a factor F = 160 compatible with 28 000 cumulative deaths, for a lockdown starting on 17 March, are assumed*.

1. *the exponential law in regime 2 is followed as long as no lockdown starts (assuming that the number of cumulative deaths is small in comparison with the population)*,
2. *the transition between the exponential-growth regime 2 and the resorption regime* 3 *is accompanied by an increase by a factor F = 160 of the cumulative death number after the lockdown*.

This extrapolation is limited by these two hypotheses, and the final numbers extracted here are either over- or under-estimated. Nevertheless, it presents the advantage to give the order of magnitude of the expected effect, and to alert on the drastic (exponential) effects of a late decision to start lockdown once the epidemics started to spread in a population.

The final number of 28 000 cumulative deaths extrapolated for a lockdown starting on 17 March would have been multiplied by a factor 7 for a lockdown started a week later, but could have been divided by the same factor 7 for a lockdown starting one week earlier. This exponential dependence of the final tolls as a function of the date of lockdown simply results from the exponential growth of deaths in regime 2.

This delay in the decision to start their lockdown measures is the main reason why the cumulative death tolls of the first epidemic waves are so high in the countries from group D, as France, Italy, Spain, the United Kingdom, the United States of America. On the contrary, the cumulative death tolls of the first epidemic waves are small in countries from group B, as Greece, Norway, Czech Republic, where lockdown was decided early, when zero or few cumulative deaths were reported.

Deviations towards smaller tolls can be expected:

i. *if a large part of the population has been contaminated (less transmissions towards non-affected cases)*
ii. *if lockdown measures are more efficient (smaller factor F)*
iii. *if local conditions slow down the mortal spread of the virus (high temperatures, low-density of population, age of the population etc.)*

On the contrary, deviations towards higher tolls can be induced by less-efficient lockdown measures (higher factor F).

### S3- What could be expected without containment measure?

Without efficient measure (mitigation, containment, lockdown), the exponential-growth regime 2 continues till saturation occurs in a natural manner, for instance if a large part of the population has been contaminated (herd immunity), or if it is slowed down by natural reasons, as perhaps a climate aggressive to the virus (temperatures, humidity etc.), a small density of population, the youth of the population etc.

Within a scenario of herd immunity, knowing that Italy and France both have approximately 60 million inhabitants, and assuming that 60 % of the population is contaminated, a fatality rate of 0.5 % (see Section 3) would lead to 200 000 cumulative deaths for each of these countries. Figure S4 shows that, without lockdown, this order of magnitude would have been approached in end of March and in beginning of April in Italy and France, respectively. Concretely, hundreds of thousands of beds equipped with respirator systems would have been needed during the epidemic peak and, due the maximal capacity to treat simultaneously a few thousands of patients, most of the patients would not have been treated, nor saved. A higher fatality rate would be reported, with perhaps a number between 500 000 and 1 million cumulative deaths.

The hypothesis of letting the virus spreading almost freely was theoretically considered as a ‘natural’ way to accept the propagation of the virus without threatening the economics of a country. This scenario of ‘herd immunity’ was initially considered in several western countries (France, United Kingdom, the Netherlands, Sweden). In the beginning of May 2020, it was abandoned in favor of a strict lockdown in all of these countries except Sweden. For this reason, a particular attention may be given to Sweden in the forthcoming weeks and months.

Oppositely, an approach aiming first to avoid high death tolls was privileged in Asian countries (South Korea, Japan, Singapore, Vietnam, Hong-Kong, Taiwan etc.), where reactive measures combined with a massive use of protection masks by the population permitted to slow down the propagation of the first epidemic waves immediately after the first confirmed cases, often without applying lockdown measures.

Countries with a high level of poverty might be unable to efficiently apply mitigation, containment, or lockdown measures to slowdown the epidemic propagation. They could in principle constitute cases where herd immunity will set up. In most of them, local specificities might constitute a natural barrier against a fast propagation of the virus.

Finally, a hasty lockdown end, or a lockdown with no appropriate measures (systematic tests on the population, massive use of masks, isolation of new confirmed cases, careful control of frontiers, quarantine for international arrivals, etc.), would lead to the resurgence of epidemic waves and to the same result, achieving herd immunity, than those expected without any lockdown.

**Figure S4:**
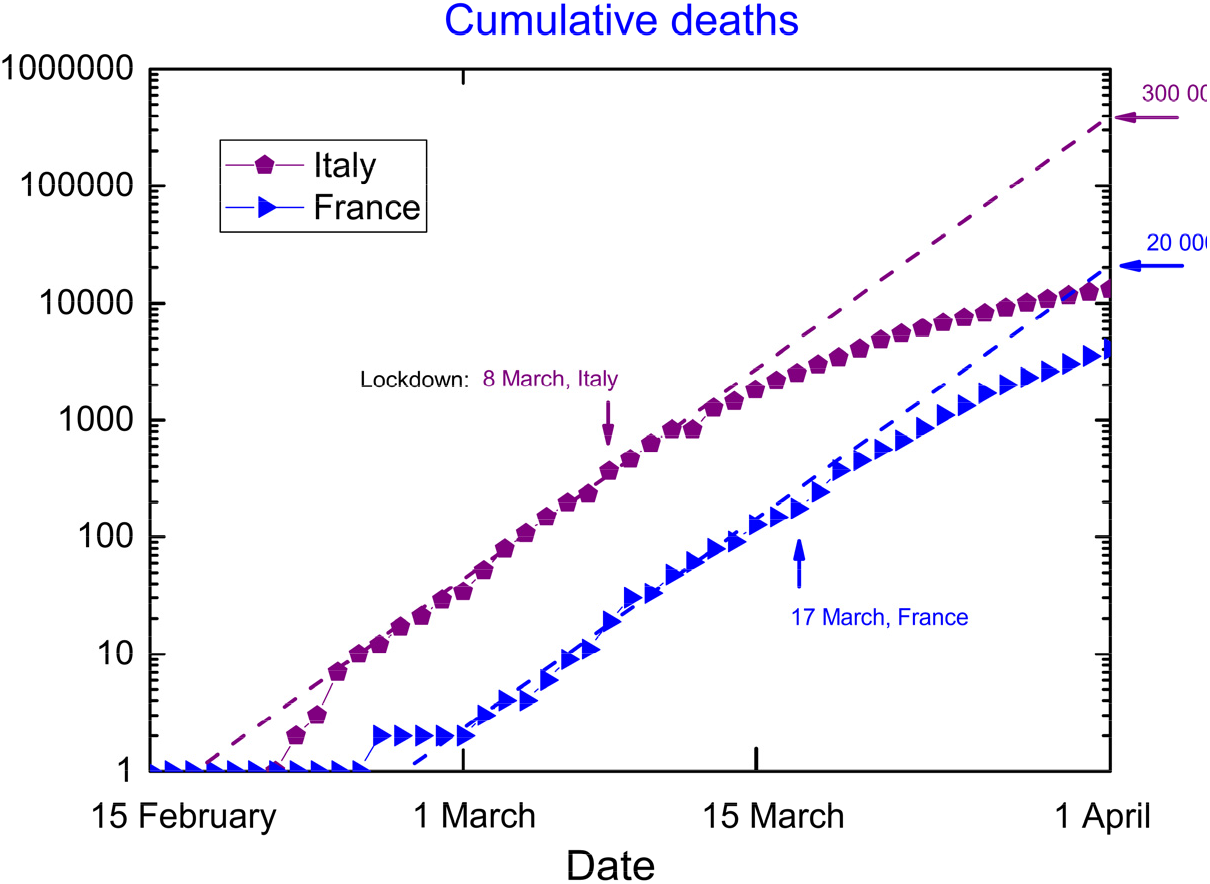
*Fit by an exponential law of the variation with time of cumulative death tolls in Italy and France, and its extrapolation after the start of lockdown*

### S4 - Focus on the propagation of COVID-19 in France

Figure S5 presents in six panels the evolution of cumulative and daily death numbers in the metropolitan regions of France, either as a function of date [Figure S5 (a-d)], or as a function of a ‘shifted’ time defined separately for each region [Figure S5(b-e)]. Cumulative and daily death numbers normalized per 100 000 inhabitants [55] are also presented [Figure S5(c-f)]. Only the deaths in the hospitals are considered in these graphs. Figure S6 presents a map of France where the shifts in time are indicated for each region. These shifts are an estimate, from data normalized with regards to the population, of the delayed start of the exponential regime 2 (see Section 3) by comparison with China.

Inhomogeneity in the epidemic propagation is observed on the French territory. The epidemic started to grow exponentially in the region Grand-Est, with a 33-days delay in comparison with China, and then expanded to the regions Ile-de-France, Hauts-de-France, and Corse, with 38-39 days delay. After these North and North-East regions, the epidemic propagated to all South-East regions and region Bretagne (West) with 40-43 days delay, before reaching all remaining regions from West and South-West with 45-46 days delay. Most of the cumulative deaths are reported in the two regions Grand Est (> 3 000 deaths) and Ile-de-France (> 6 000 deaths), which were hit first.

For all regions, the same exponential-growth regime is observed at the beginning of the epidemic propagation. Lockdown has been applied to the whole country on 17 March and a deviation from the exponential growth of the death tolls can be seen ten days later in most regions. After the 15 April a decay of the daily death tolls is observed in all regions. In region Ile-de-France, an upward variation is visible a few days after the lockdown start, and a downturn deviation from the exponential growth appeared only two weeks later. Without surprise, the lower death tolls are reported in the regions where the epidemic arrived later. In these West and South-West regions, lockdown started when the death tolls were low. It permitted to contain their increase more efficiently than in the North and North-East regions. Disparity between regions is evidenced in Figure S5(c,f), where the death tolls / 100 000 inhabitants are shown to be 20 times higher in regions Grand-Est and Ile-de-France than in the less-impacted regions. Region Ile-de-France follows region Grand-Est with 5 days delay, indicating that lockdown start has been less efficient in Ile-de-France.

Figure S7 presents the evolution of the death tolls in a selection of the mostly-impacted French departments. Only deaths in hospitals are considered in these graphs. Figure S8 shows a map of the East of France where the shifts in time are indicated for each of the considered departments. These shifts were defined from the numbers of deaths normalized per 100 000 inhabitants [56]. They give an estimation of the delay of the exponential regime 2 start, by comparison with China [Figure S7(c-f)]. After the 15 April a decline of the daily death tolls is observed in all considered departments. These graphs confirm that the deviation from regime 2 occurs later in departments from region Ile-de-France than in the other departments. A maximum of 4 daily deaths per 100 000 inhabitants has been reported in the department Bas-Rhin and corresponds to the maximal value observed in Belgium, the highest national maximum reported in the world before mid-May 2020.

**Figure S5:**
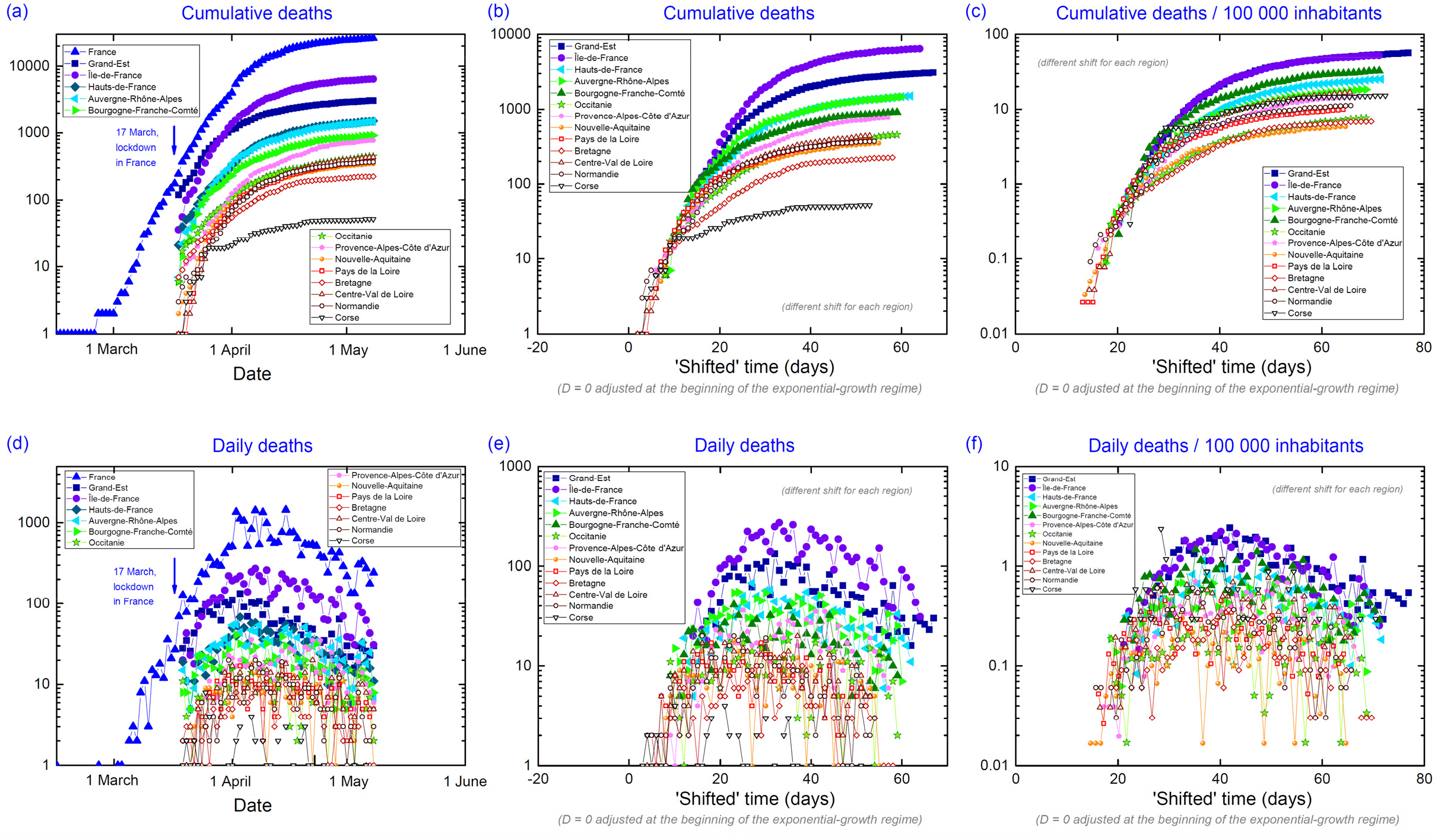
*Focus on cumulative and daily deaths in French metropolitan regions*.

**Figure S6:**
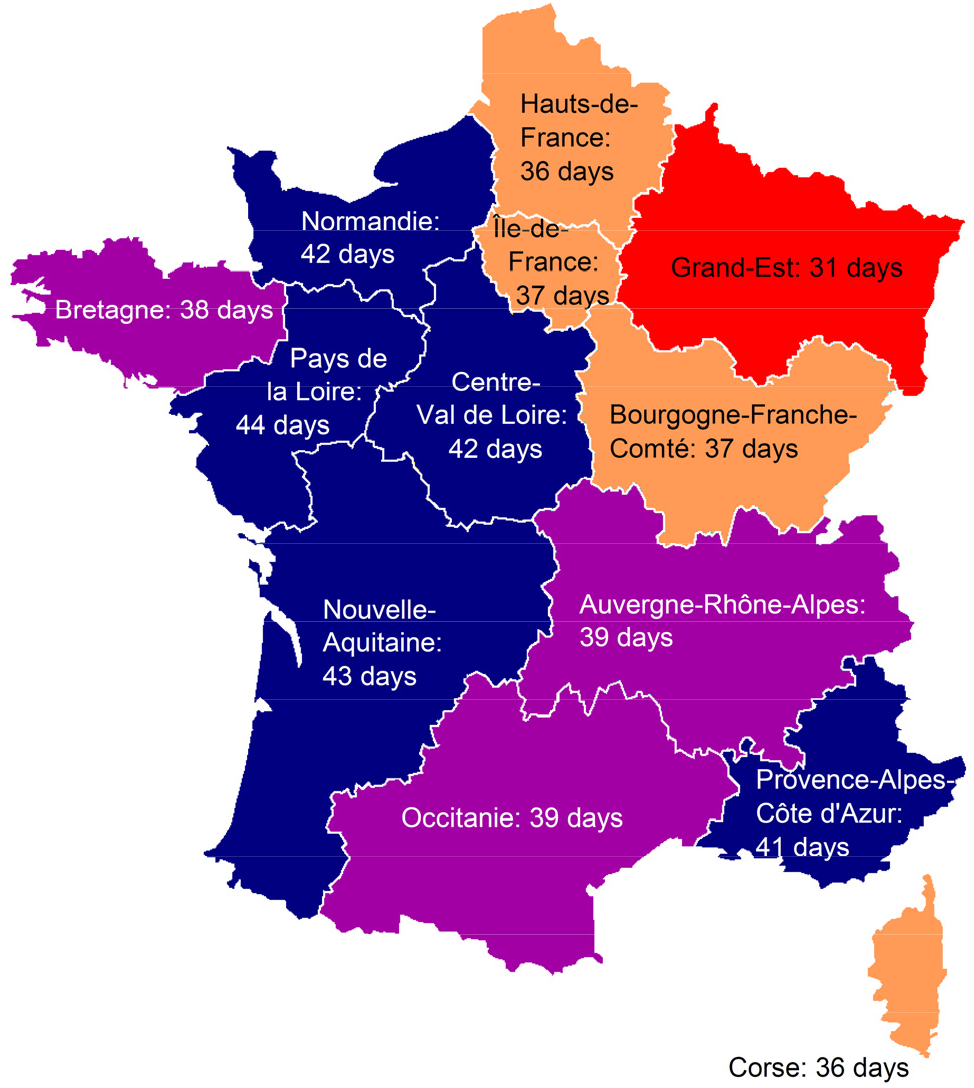
*Map of France and shift in days of the beginning of the epidemic exponential-growth regime 2 for the metropolitan regions in comparison with China*.

**Figure S7:**
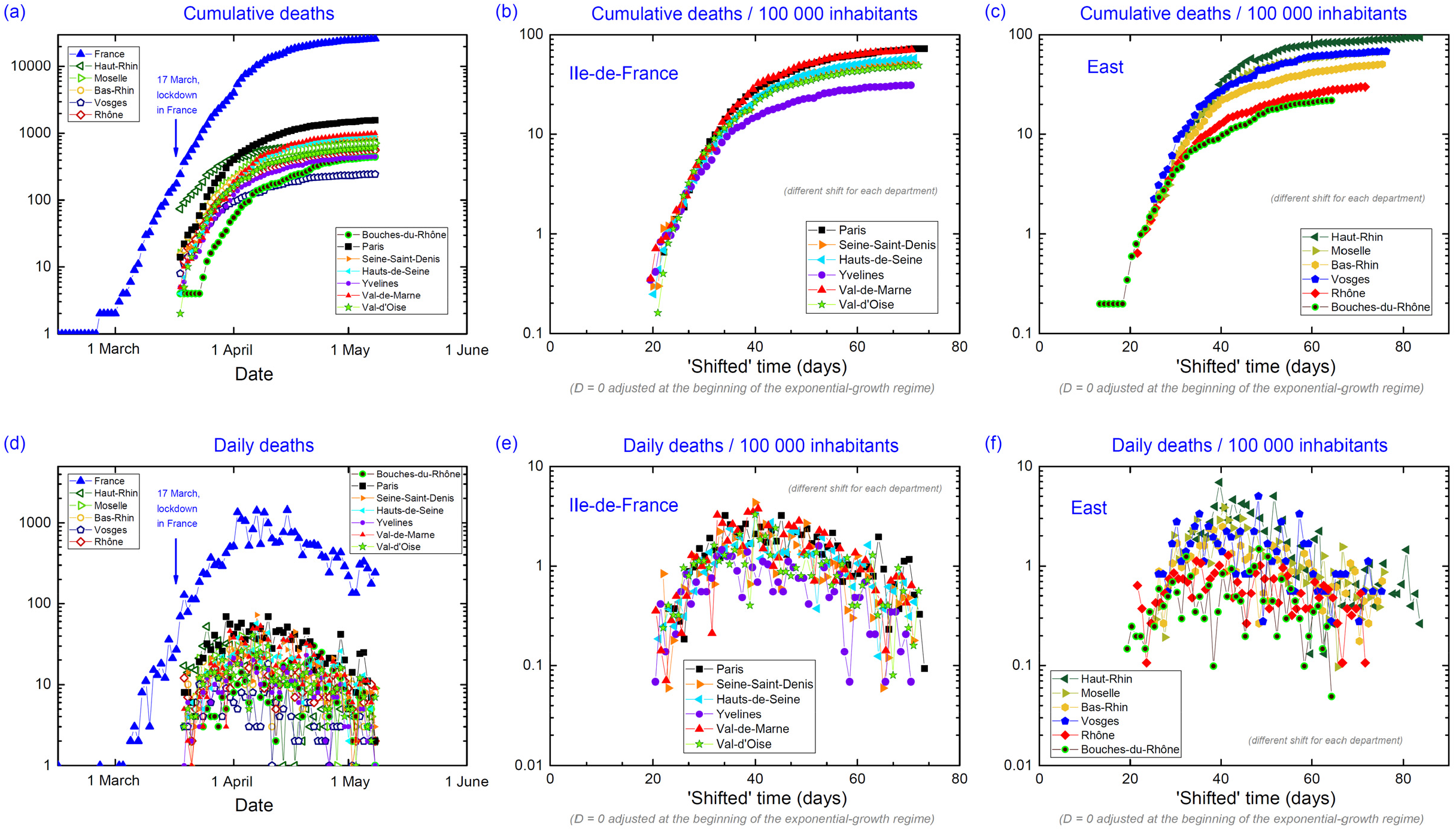
*Cumulative deaths, daily deaths, and normalized daily deaths per 100 000 inhabitants as function of date or ‘shifted’ time for a selection of French departments in region Ile-de-France and in the East of the country*.

**Figure S8:**
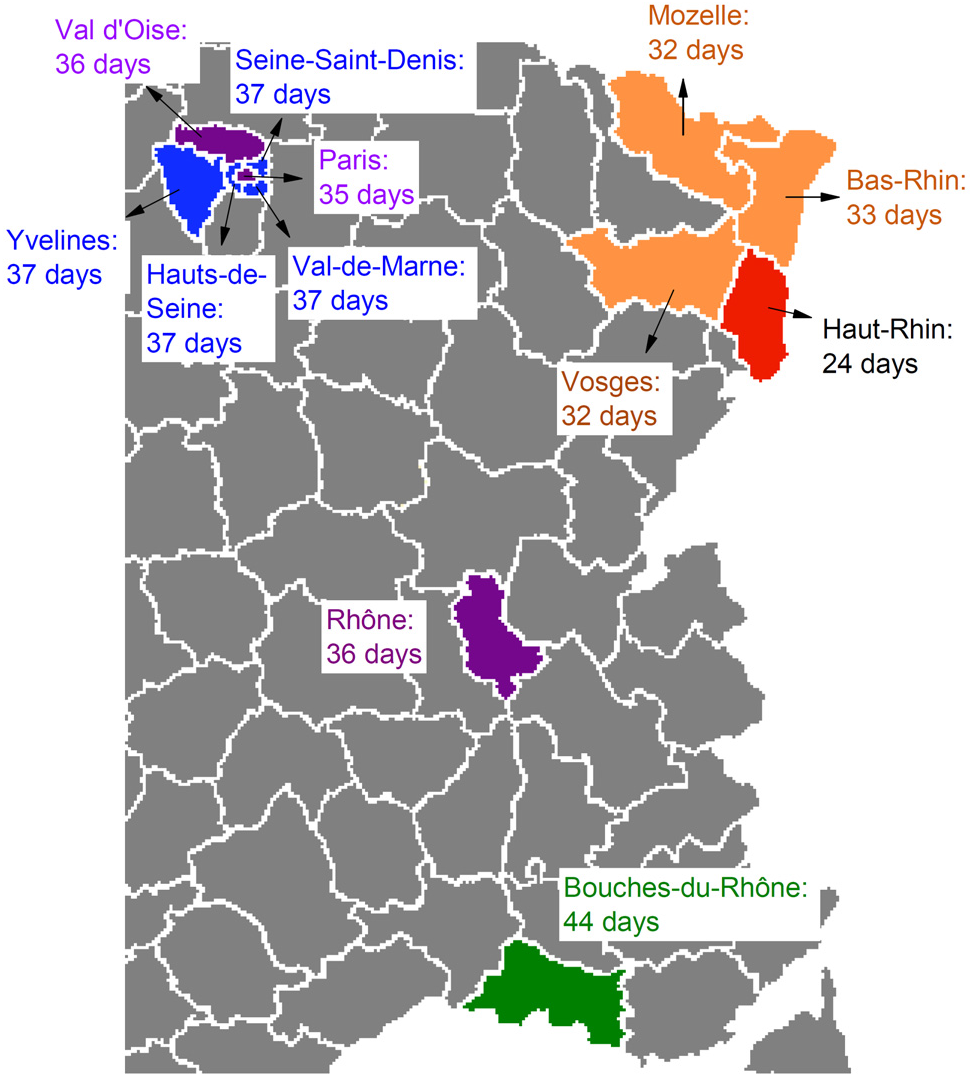
*Map of East of France and shift in days at the beginning of the epidemic exponential-growth regime 2 for a selection of departments, in comparison with China*.

Back to a national scale, we can briefly discuss the political measures and their effects in France. Figure S9 presents the evolution of cumulative and daily death tolls in France, with annotations indicating political recommendations and measures, as well as the last observed mass events. A downturn deviation from exponential growth regime was observed on 15 March, but it has been almost cancelled by a 50 % daily increase of the cumulative deaths on 19 March. National lockdown was applied on 17 March. After the 2 April, the deaths in nursing homes were also counted, leading to an upward correction of the cumulative death toll. Figure S9(b) indicates that the epidemic peak can be identified on 11 April, in the center of a 20-days long plateau in the daily death toll. The decline of daily deaths observed in the second half of April is the consequence of the lockdown measures taken three weeks before. Figure S8 shows no evidence that the different and progressive measures taken from 5 to 17 March had some effect against the pandemic propagation. As detailed in Section S2, a simple consequence of these progressive measures was a delay in the application of strong and efficient measures, which could have led to a much smaller death toll if applied earlier.

**Figure S9:**
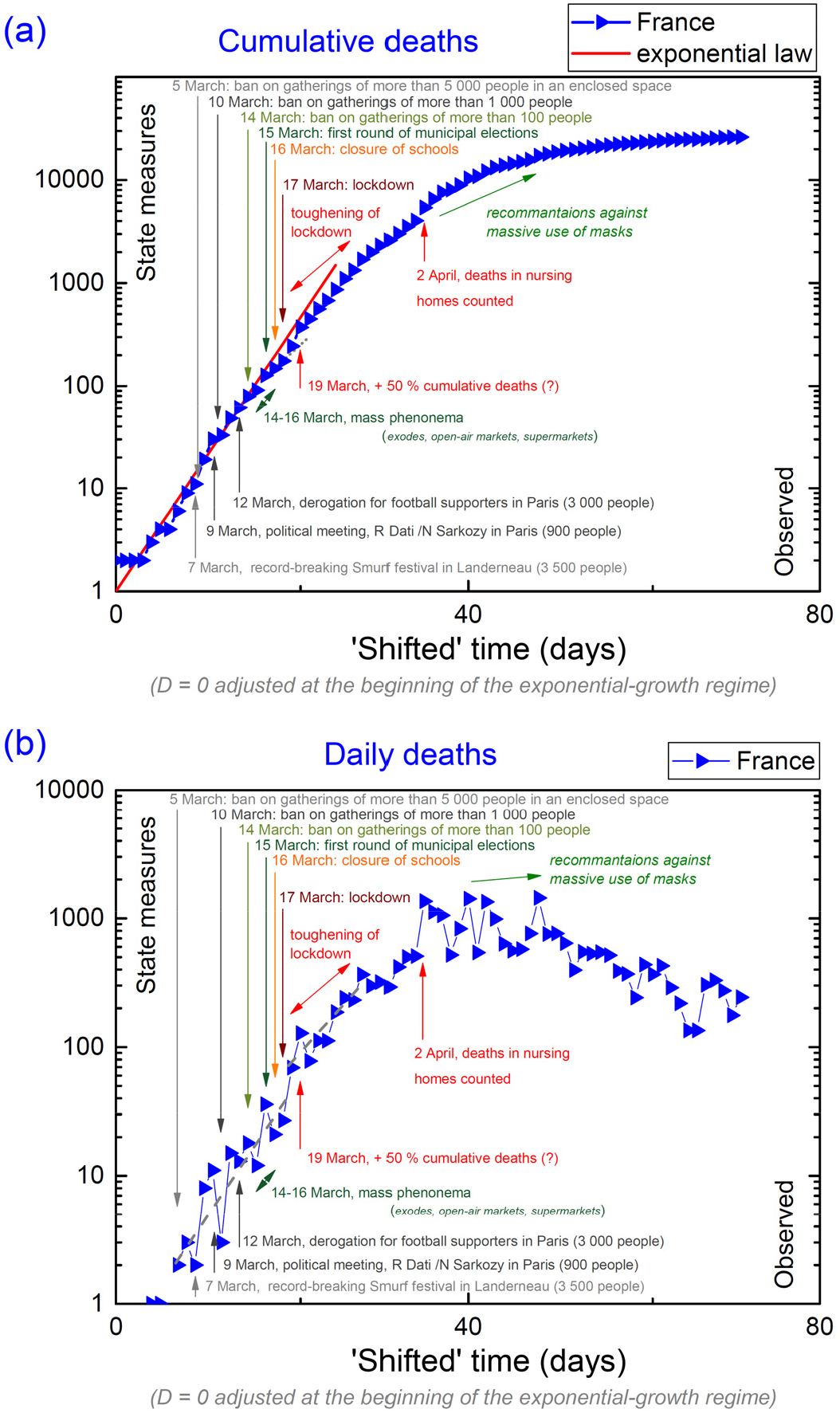
*Variation of (a) cumulative and (b) daily death tolls in France, in regards with political measures and the last mass events*.

## Notes

### Competing Interest Statement

The authors have declared no competing interest.

### Funding Statement

no external funding

